# Improved Prediction of COVID-19 Transmission and Mortality Using Google Search Trends for Symptoms in the United States

**DOI:** 10.1101/2021.03.14.21253554

**Authors:** Meshrif Alruily, Mohamed Ezz, Ayman Mohamed Mostafa, Nacim Yanes, Mostafa Abbas, Yasser EL-Manzalawy

## Abstract

Accurate forecasting of emerging infectious diseases can guide public health officials in making appropriate decisions related to the allocation of public health resources. Due to the exponential spread of the COVID-19 infection worldwide, several computational models for forecasting the transmission and mortality rates of COVID-19 have been proposed in the literature. To accelerate scientific and public health insights into the spread and impact of COVID-19, Google released the Google COVID-19 search trends symptoms open-access dataset. Our objective is to develop 7 and 14 -day-ahead forecasting models of COVID-19 transmission and mortality in the US using the Google search trends for COVID-19 related symptoms. Specifically, we propose a stacked long short-term memory (SLSTM) architecture for predicting COVID-19 confirmed and death cases using historical time series data combined with auxiliary time series data from the Google COVID-19 search trends symptoms dataset. Considering the SLSTM networks trained using historical data only as the base models, our base models for 7 and 14 -day-ahead forecasting of COVID cases had the mean absolute percentage error (MAPE) values of 6.6% and 8.8%, respectively. On the other side, our proposed models had improved MAPE values of 3.2% and 5.6%, respectively. For 7 and 14 -day-ahead forecasting of COVID-19 deaths, the MAPE values of the base models were 4.8% and 11.4%, while the improved MAPE values of our proposed models were 4.7% and 7.8%, respectively. We found that the Google search trends for “pneumonia,” “shortness of breath,” and “fever” are the most informative search trends for predicting COVID-19 transmission. We also found that the search trends for “hypoxia” and “fever” were the most informative trends for forecasting COVID-19 mortality.

## Introduction

In March 1^st^, 2020, the COVID-19 outbreak was declared a national emergency in the US. After exactly one year of this declaration and according to the JHU dashboard, the numbers of COVID-19 confirmed and death cases have reached more than 500K and 17M, respectively. This rapid spread of the virus in the US had negative impacts on several sectors including economy [1], education [2-4], health [5, 6]. Reliable real-time forecasting of the spread of infectious diseases, including COVID-19, can improve public health response to outbreaks and save lives [7, 8].

Since the emergence of the COVID-19 outbreak in late 2019, several scientists have developed computational models for forecasting COVID-19 confirmed cases, deaths, and recovery [9, 10]. Commonly used statistical methods for time series forecasting such as autoregressive integrated moving average (ARIMA) [11] have been used in multiple studies for forecasting COVID-19 (e.g., [12-14]). These methods are typically based on historical data and do not account directly for disease transmission dynamics or any relevant biological process [15-17]. To account for these factors, epidemiological methods have been proposed for forecasting infectious diseases. Examples of the application of epidemiological methods for modeling the spread of COVID-19 infection include several frameworks based on the adaption of the SEIR (Susceptible, Exposed, Infected, Recovered) method [18-20]. Because the time series forecasting task can be formulated as a supervised learning problem [21], several machine learning algorithms have been used for forecasting COVID-19 (e.g., [22-25]).

Recurrent neural networks (RNNs) [26, 27] are machine learning based models that have been successfully applied to the problem of forecasting time series [28-30]. In RNNs, recurrent layers consist of a sequence of recurrent cells whose states are determined by past states and current inputs. Long short-term memory (LSTM) [31, 32] is a variant of RNN designed to capture long-term dependencies by introducing gate functions into the recurrent cell structure. Since its introduction, LSTM is probably the most widely used form of RNNs and have been successfully used in a broad range of sequence classification tasks including financial time series prediction [33], speech recognition [34], sentiment classification [35], traffic forecasting [36], and anomaly detection [37]. Deep neural network architectures can better model real-world time series with complex non-linear relationships [38, 39]. A deep LSTM architecture, also called stacked LSTM (SLSTM), consists of several hidden LSTM layers and has been shown more effective in modeling complex sequence data [40].

Recently, Abbas et al. [41] have shown that Google search trends for nine COVID-19 related symptoms (namely, hypoxemia, ageusia, anosmia, dysgeusia, hypoxia, fever, pneumonia, chills, and shortness of breath (SOB)) are strongly associated with COVID-19 confirmed as well as death cases in the US. Results of their analysis suggested that these Google trends can be used (in combination with COVID-19 historical data) to forecast COVID-19 spread and mortality up to three weeks ahead in time. The main goal of this study is to validate this finding. Specifically, we propose a stacked LSTM (SLSTM) model for forecasting state-level daily cumulative COVID-19 confirmed and death cases in the US. We then use this model to quantify the importance of each Google search trend for forecasting COVID-19 transmission and mortality in the US. Finally, we demonstrate substantial improvements in the predictive performance of the SLTSM models when the data for up to three Google search trends are incorporated in the training of these models. Our results demonstrate the viability of incorporating Google search trends for COVID-19 related symptoms into deep learning models for forecasting COVID-19 transmission and mortality in the US.

## Methods

### Data

Daily cumulative counts for COVID-19 confirmed and death cases were downloaded from a publicly available repository maintained by the Center for Systems Science and Engineering (CSSE) at Johns Hopkins University (JHU) [42]. We considered the data aggregated at the state level for the 50 US states plus the District of Columbia. We experimented with the data from March 1^st^, 2020 to September 31^st^, 2020. The downloaded counts were then normalized to count per million people in each state using 2019 census population estimates.

State-level aggregated and normalized Google COVID-19 search trends symptoms [43] were downloaded from https://github.com/google-research/open-covid-19-data/. The data includes search trends for 422 symptoms that might be related to COVID-19. However, we limited our experiments to the nine symptoms suggested by the exploratory functional data analysis [44] provided in [41]. Therefore, our final symptoms dataset includes time series for the following nine symptoms: hypoxemia, ageusia, anosmia, dysgeusia, hypoxia, fever, pneumonia, chills, and shortness of breath (SOB).

Each time series were split into three sets for training, validation, and testing. The test set covers the last 45 days in our study interval (i.e., from August 17^th^ to September 31^st^). The data for the remaining study time (from March 1^st^ to August 16^th^) were split into training and validation such that the data from the last 45 days in this interval were used for validation.

### *k*-day-ahead forecasting

Given a time series with *n* time points, *x*_1_, …., *x*_*n*_, the goal is to predict time series at the future time points (i.e., *n* + 1, *n* + 2, etc.). For *k*-step-ahead forecasting, the predictive model is required to predict *x*_*n*+*k*_ given historical data up to *x*_*n*_. Often, the predictive model does not use the entire historical data but only uses the *w* most recent time points (e.g., *x*_*n*-*w*+1_, …, *x*_*n*_). These fixed-length windows labeled with the target outcome *y* = *x*_*n*+*k*_ are ideal for training machine learning models. In this study, since every step in the time series is a day, we call the *k*-step-ahead forecasting a *k*-day-ahead forecasting.

In the presence of an auxiliary time series (i.e., one time series corresponding to the Google search trends for COVID-19 related symptoms), *s*_1_, …, *s*_*n*_, we can generate labeled samples in two spaces: i) symptom space, < [*s*_*n*-*w*+1_, …, *s*_*n*_], *x*_*n*+1_ >; ii) historical + symptom space,

< [*x*_*n*-*w*+1_, …, *x*_*n*_, …, *s*_*n*-*w*+1_, …, *s*_*n*_], *x*_*n*+1_ >. This can also be generalized to the case when multiple auxiliary time series are available.

### Our proposed deep learning model

We used the Long short-term memory (LSTM) networks [31, 32] for developing predictive models for the four COVID-19 forecasting tasks considered in this study. LSTM is a type of Recurrent Neural Network (RNN) that is suitable for learning long-term dependencies in sequence data [45]. Long-terms dependencies are modeled using memory blocks [46]. A memory block or a single LSTM cell is a recurrently connected sub-network that contains a memory cell and three gates. Fig. 1 shows the architectures of an LSTM cell in an LSTM layer. The memory cell remembers the temporal state of the cell and the gates control the pattern of information flow. Input and output gates control information flow into and from the cell, respectively. The forget gate controls what information will be thrown away from the memory cell. Mathematically, an LSTM is expressed using the following equations:

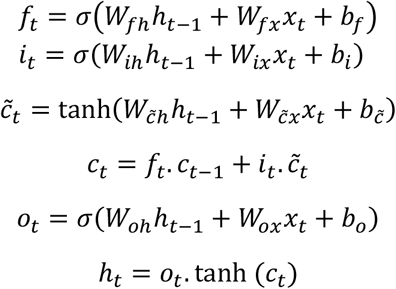

**Figure 1.**
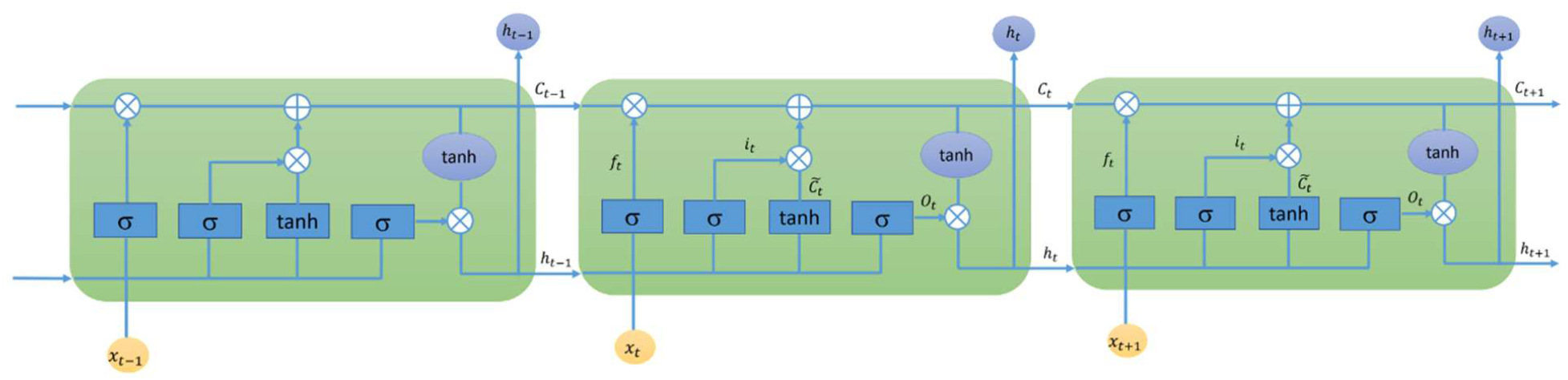
LSTM Layer.

A simple LSTM network includes a single LSTM layer. To add capacity and depth, multiple LSTM layers could be stacked together to form a multilayer fully connected structure [47]. Fig. 2 shows the structure of our proposed stacked LSTM (SLSTM) network, which included two LSTM layers. The first and second LSTM layers had 128 and 64 hidden units, respectively. The output of the second LSTM layer represents the deep features learned from the sequence data, which is then fed to a dense layer with 64 units followed by a single neuron, fully connected to the 64 neurons from the previous layer, for output. It is worth noting that this architecture had been used across all forecasting tasks and data representations as shown in the following subsection.

**Figure 2.**
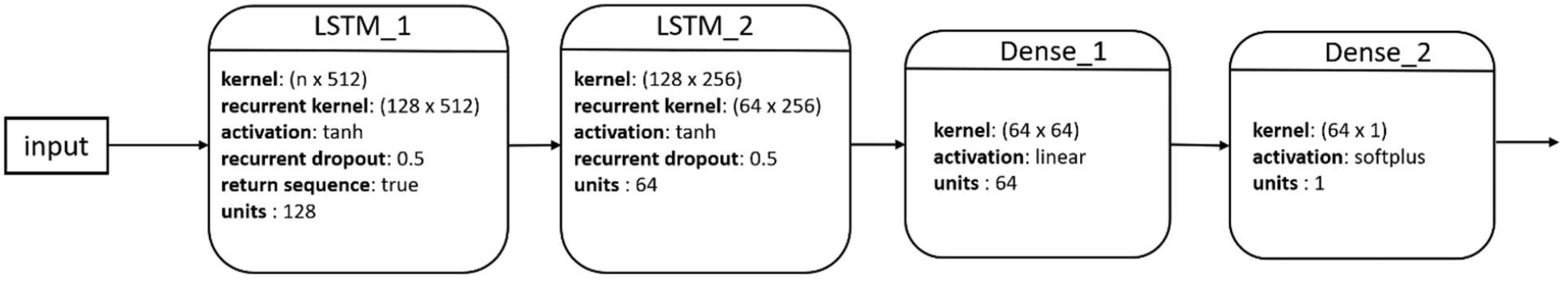
Architecture of our proposed SLSTM model.

### Experimental settings

In our experiments, we considered *k*-day-ahead forecasting of COVID-19 confirmed cases for *k* = 7 and 14 days. We also experimented with *k*-day-ahead forecasting of COVID-19 death cases for *k* = 7 and 14 days. For these four prediction tasks, we experimented with the following types of input features: i) historical data; ii) individual Google search trend for nine COVID-19 related symptoms; iii) historical data and single Google search trend for nine COVID-19 related symptoms; iv) historical data and top two Google search trends for nine COVID-19 related symptoms determined using the validation set in step 3 experiments; v) historical data and top three Google search trends for nine COVID-19 related symptoms determined using the validation set in step 3 experiments. Hence, the number of input features ranged from *w* to *w* + *w* × *d*, where *w* is the window size parameter and *d* = {0,1,2,3} is the number of auxiliary time series used. Because the optimal window size, *w*, is often data and task -dependent, we experimented with *w* = {2,3, …, 9} and determined the optimal value for *w* using the performance of the learned model on the validation set.

For implementing and evaluating the stacked LSTM models, we used the Keras version 2.4.3 and tensorflow version 2.3.1 libraries. For all experiments resulting from all possible combinations of the four forecasting tasks and the five types of inputs, we used the architecture and configurations shown in Fig. 2. The activation functions used were *tanh, linear*, and *softplus* for the two LSTM layers, dense layer, and output neuron, respectively. For the two LSTM layers, the recurrent dropout rate was set to 0.5. For training our models, we used the Mean Squared Logarithmic Error (MSLE) loss and an early stopping technique [48] such that the training process was stopped if no improvement in the model performance, in terms of MAPE on the validation set, was noted for 20 iterations.

We assessed the predictive performance of our models using the mean absolute percentage error (MAPE) defined as 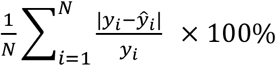, where *N* is the number of time points in the test time series, *y*_*i*_ is the *i*^th^ target outcome, and *ŷ*_*i*_ is the *i*^th^ predicted outcome.

## Results

### Trajectories of COVID-19 confirmed and death cases

Fig. 3 shows the cumulative COVID-19 confirmed case (left) and death (right) trajectories for the 51 US states. For COVID-19 confirmed cases, we noted that NY and NJ had the largest total per million counts of confirmed COVID-19 cases during mid-March until the third week of July. Starting the third week of July, several states, including LA, FL, and AZ, exceeded the number of confirmed cases in NY and NY as the rates of COVID-19 spread started to drop substantially in these two states. For COVID-19 death cases, we found that NJ and NY consistently had the highest number of total deaths and that their curves seemed to be flat starting the third week of July.

**Figure 3.**
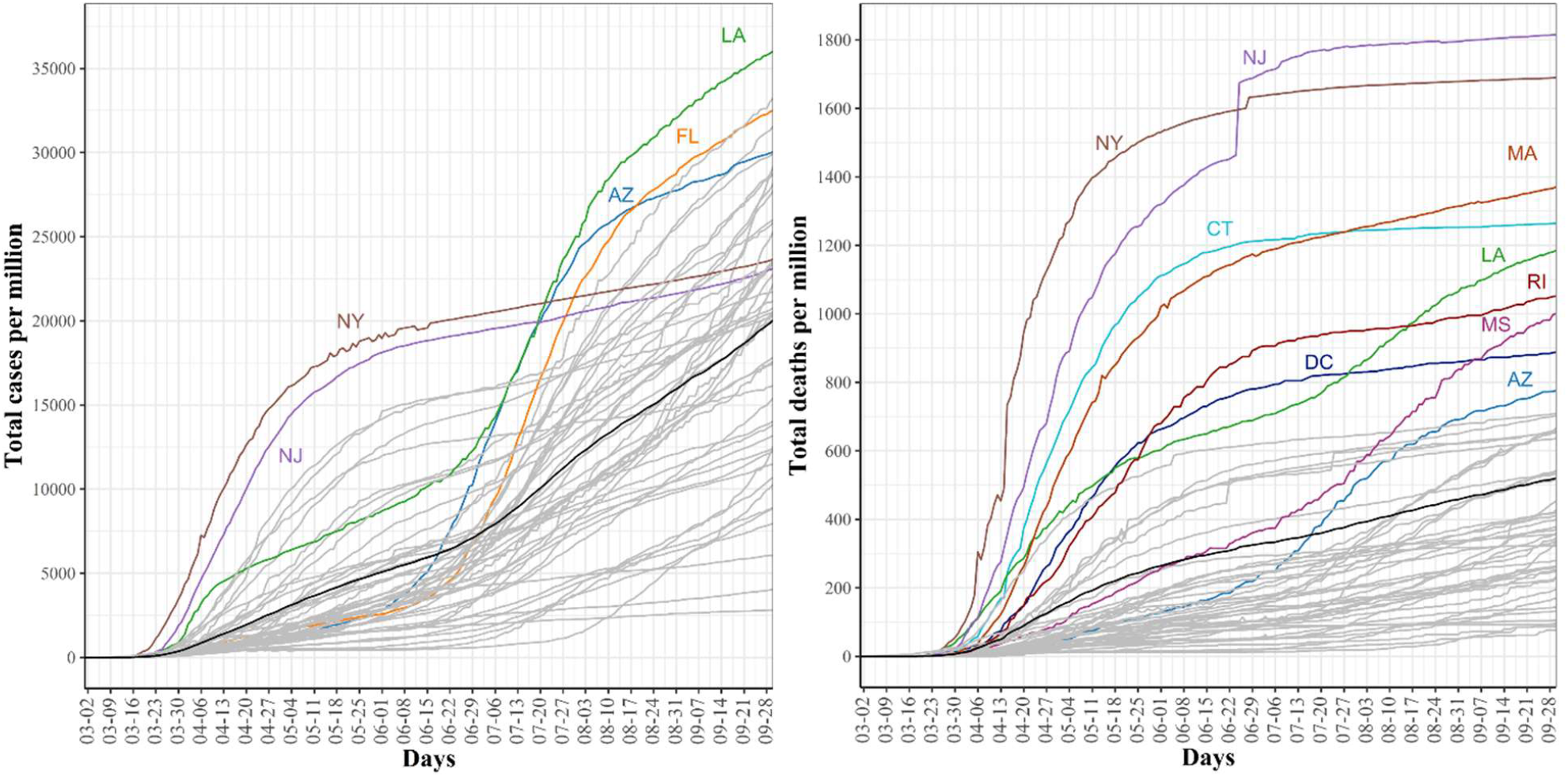
Trajectories for COVID-19 daily cumulative per million confirmed (left) and death (right) cases. The pointwise mean curve is highlighted in black.

### Prediction of COVID-19 confirmed and death cases using historical data and univariate SLSTM

We report the performance of SLSTM models for predicting COVID-19 cases and mortality *k*-day-ahead for *k* equals 7 and 14 days. Table 1 shows the performance of 16 LSTM models for forecasting COVID-19 cases. For the 7-day-ahead COVID-19 case prediction, we found that the best model determined using its MAPE score on the validation set has MAPE scores of 7.4% and 6.6% on validation and test sets, respectively. This model used a window of size equals 9. We also noted that a better MAPE score of 5.5% was obtained using a window of size equals 5, but its performance on the validation set did not recommend selecting it as the optimal learned model. For 14-day-ahead COVID-19 case prediction, the best performing model used a window of size equals 9 and had the best observed MAPE scores of 11.8% and 8.8% on validation and test sets, respectively.

**Table 1:** Performance (in terms of MAPE) of different SLSTM for forecasting COVID-19 confirmed cases using historical data only and different window sizes.

| Window Size | 7-day-ahead |  | 14-day ahead |  |
| --- | --- | --- | --- | --- |
|  | Validation | Test | Validation | Test |
| 2 | 9.4 | 8.8 | 13.5 | 11.6 |
| 3 | 9.8 | 7.8 | 13.7 | 11.6 |
| 4 | 10.1 | 7.7 | 13.9 | 12.6 |
| 5 | 7.9 | **5.5** | 14.2 | 11.9 |
| 6 | 7.9 | 6.3 | 13.1 | 9.8 |
| 7 | 8.0 | 6.4 | 14.5 | 11.1 |
| 8 | 8.3 | 7.1 | 12.7 | 9.1 |
| **9** | **7.4** | **6.6** | **11.8** | **8.8** |

Table 2 shows the performance of 16 SLSTM models for forecasting COVID-19 death cases. For the 7-day-ahead COVID-19 death prediction, the best SLSTM model was obtained using a window of size equals 6 and had the best observed MAPE scores of 7.3% and 4.8% on the validation and test sets, respectively. For the 14-day-ahead COVID-19 death prediction, the best model used a window of size equals 7 and had the lowest noted MAPE scores of 13.1% and 11.4% on the validation and test sets, respectively.

**Table 2:** Performance (in terms of MAPE) of different SLSTM for forecasting COVID-19 death cases using historical data only and different window sizes.

| Window Size | 7-day ahead |  | 14-day ahead |  |
| --- | --- | --- | --- | --- |
|  | Validation | Test | Validation | Test |
| 2 | 24.1 | 13.0 | 27.2 | 14.4 |
| 3 | 24.8 | 14.2 | 26.7 | 14.5 |
| 4 | 11.5 | 9.1 | 19.3 | 15 |
| 5 | 9.4 | 7.2 | 19.6 | 14.3 |
| **6** | **7.3** | **4.8** | 21.2 | 13.6 |
| **7** | 10.3 | 8.0 | **13.1** | **11.4** |
| 8 | 100 | 100.0 | 99.9 | 100 |
| 9 | 100 | 100.0 | 100 | 100 |

Interestingly, we found that: i) MAPE scores for best performing COVID-19 case prediction models were better than those for the best performing COVID-19 death prediction models; ii) Performance of the 7-day-ahead forecasting models was better than the performance of the 14-day-ahead forecasting models; iii) Performance of the models estimated using the validation sets was consistently lower than the performance of the models estimated using the test set. However, the validation test successfully identified the best performing model on the test set for 3 out of four prediction tasks.

### Improved prediction of COVID-19 confirmed and death cases using Google search trends for COVID-19 symptoms

We proceed with reporting our experimental results for testing two hypotheses regarding the Google search trends for nine COVID-19 related symptoms: i) SLSTM models trained using any single symptom can predict COVID-19 confirmed and death cases; ii) SLSTM models trained using any single symptom combined with COVID-19 historical data can better predict COVID-19 confirmed and death cases compared with SLSTM models trained using historical data only. Our rationale is that these nine symptoms had been shown to have strong associations with both of COVID-19 transmission and mortality [41] and, therefore, could be used for forecasting COVID-19 transmission and mortality or for improving the performance of the models developed for the four tasks considered in this study.

Tables 3 and 4 show that the SLSTM models based on any of these nine symptoms failed to accurately predict COVID-19 cases 7 and 14 -day-ahead. For these models, the best observed MAPE score was around 60%. However, when any of these symptoms were combined with historical COVID-19 cases data, the best performing models, identified using the validation set, had MAPE scores on the test that was lower than the MAPE scores of the best performing SLSTM models trained using historical data only (i.e., MAPE scores of 6.6% and 8.8% for forecasting COVID-19 cases 7 and 14 -day-ahead, respectively). We also noted that, for predicting COVID-19 cases 7-day-ahead, the optimal SLSTM model used a window of size equals 3 and a combination of pneumonia and historical data as input. On the other hand, for predicting COVID-19 cases 14-day-ahead, the optimal SLSTM model used a window of size equals 5 and a combination of pneumonia and historical data as input. Despite these performance improvements, we observed that the MAPE scores estimated using the validation set failed to identify the best performing model on the test set. Overall, our results rejected the first hypothesis and accepted the second one for forecasting COVID-19 cases.

**Table 3:**
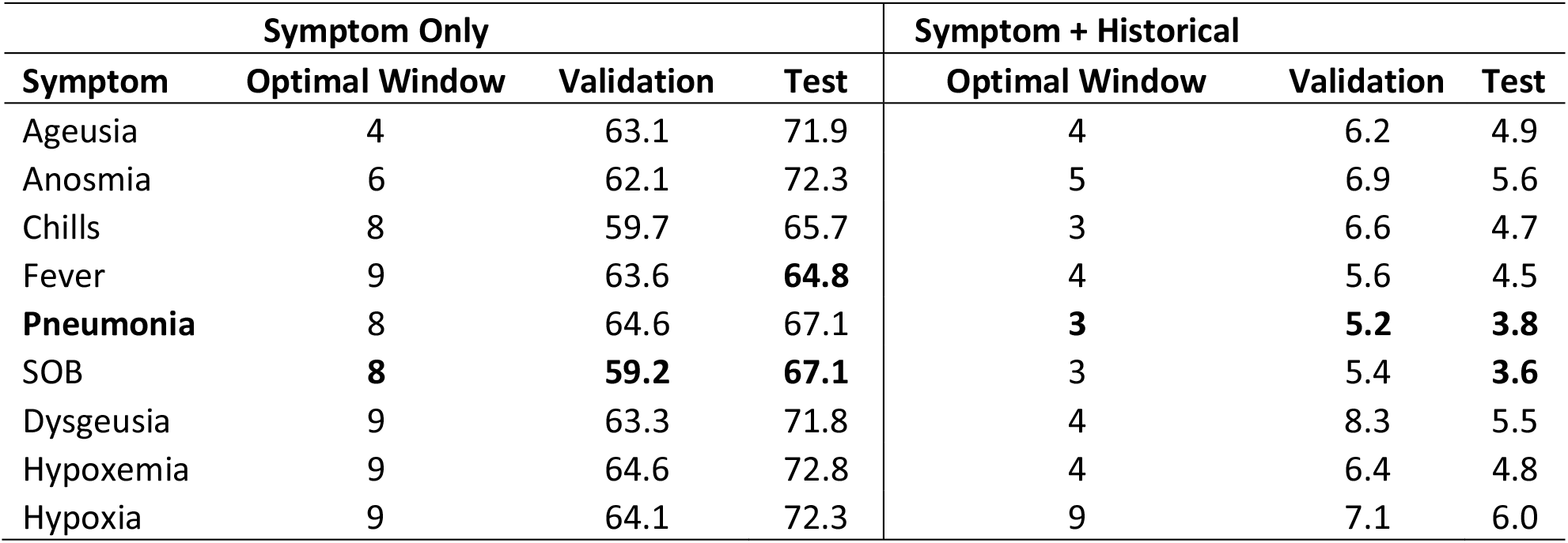
Performance of optimal SLSTM models for 7-day-ahead forecasting of COVID-19 confirmed cases when the models were trained using a single symptom time series alone or combined with historical data.

| Symptom Only |  |  |  | Symptom + Historical |  |  |
| --- | --- | --- | --- | --- | --- | --- |
| Symptom | Optimal Window | Validation | Test | Optimal Window | Validation | Test |
| Ageusia | 4 | 63.1 | 71.9 | 4 | 6.2 | 4.9 |
| Anosmia | 6 | 62.1 | 72.3 | 5 | 6.9 | 5.6 |
| Chills | 8 | 59.7 | 65.7 | 3 | 6.6 | 4.7 |
| Fever | 9 | 63.6 | **64.8** | 4 | 5.6 | 4.5 |
| **Pneumonia** | 8 | 64.6 | 67.1 | **3** | **5.2** | **3.8** |
| SOB | **8** | **59.2** | **67.1** | 3 | 5.4 | **3.6** |
| Dysgeusia | 9 | 63.3 | 71.8 | 4 | 8.3 | 5.5 |
| Hypoxemia | 9 | 64.6 | 72.8 | 4 | 6.4 | 4.8 |
| Hypoxia | 9 | 64.1 | 72.3 | 9 | 7.1 | 6.0 |

**Table 4:** Performance of optimal SLSTM models for 14-day-ahead forecasting of COVID-19 confirmed cases when the models were trained using a single symptom time series alone or combined with historical data.

| Symptom Only |  |  |  | Symptom + Historical |  |  |
| --- | --- | --- | --- | --- | --- | --- |
| Symptom | Optimal Window | Validation (rank) | Test | Optimal Window | Validation (rank) | Test |
| Ageusia | 7 | 62.2 (3.0) | 70.0 | 4 | 10.7 (4.0) | 8.1 |
| Anosmia | **9** | **61.4 (1.5)** | **71.2** | 5 | 11.2 (5.0) | 8.7 |
| Chills | **7** | **61.4 (1.5)** | **66.6** | 3 | 11.9 (8.0) | 8.1 |
| Fever | 3 | 62.7 (5.0) | 67.3 | 3 | 9.7 (2.0) | 6.5 |
| **Pneumonia** | 3 | 63.5 (8.0) | 65.3 | **5** | **9.6 (1.0)** | **7.4** |
| SOB | 3 | 62.4 (4.0) | 67.1 | 3 | 10.0 (3.0) | **6.1** |
| Dysgeusia | 6 | 62.9 (7.0) | 71.6 | 7 | 14.4 (9.0) | 9.6 |
| Hypoxemia | 7 | 64.4 (9.0) | 72.5 | 7 | 11.6 (6.0) | 8.2 |
| Hypoxia | 3 | 62.8 (6.0) | 71.0 | 9 | 11.8 (7.0) | 9.3 |

Similar findings were found for forecasting COVID-19 deaths (See Tables 5 and 6). Using any single symptom for training the SLSTM models for predicting COVID-19 death 7 and 14 -day-ahead yielded poor models with MAPE scores of 75% or higher. When using search trends for hypoxia combined with historical death counts, the best SLSTM had a MAPE score of 4.7%. That was a slight performance improvement over the best SLSTM model using historical death counts only with MAPE score of 4.8% for predicting COVID-19 deaths 7-day-ahead. However, there existed a model with an improved MAPES score of 3.9%, but our selection criteria of the best model based on its performance on the validation set failed to suggest it. For 14-day-ahead forecasting of COVID-19 deaths, the best SLSTM model, based on search trends for favor and historical death counts, had a MAPE score of 8.7%, which is a considerable performance improvement compared with the SLSTM model based on historical data only with a MAPE score of 11.4%.

**Table 5:** Performance of optimal SLSTM models for 7-day-ahead forecasting of COVID-19 death cases when the models were trained using a single symptom time series alone or combined with historical data.

| Symptom Only |  |  |  | Symptom + Historical |  |  |
| --- | --- | --- | --- | --- | --- | --- |
| Symptom | Optimal Window | Validation (rank) | Test | Optimal Window | Validation (rank) | Test |
| Ageusia | 8 | 77.5 (3.0) | 57.3 | 6 | 15.4 (9.0) | 9.6 |
| Anosmia | 8 | **75.0 (1.5)** | **56.4** | 6 | 11.4 (7.0) | 6.6 |
| Chills | 8 | 87.7 (6.0) | 57.9 | 7 | 7.8 (2.0) | 6.2 |
| Fever | 4 | 112.7 (7.0) | 69.9 | 6 | 8.4 (4.0) | **3.9** |
| Pneumonia | 3 | 129.2 (9.0) | 72.5 | 7 | 9.2 (5.0) | 6.1 |
| SOB | 3 | 112.8 (8.0) | 70.5 | 7 | 8.3 (3.0) | 5.2 |
| Dysgeusia | 3 | **75.0 (1.5)** | **54.7** | 6 | 12.6 (8.0) | 10.3 |
| Hypoxemia | 3 | 80.8 (5.0) | 60.1 | 7 | 9.7 (6.0) | 6.3 |
| **Hypoxia** | 5 | 78.7 (4.0) | 60.4 | **6** | **6.7 (1.0)** | **4.7** |

**Table 6:** Performance of optimal SLSTM models for 14-day-ahead forecasting of COVID-19 death cases when the models were trained using a single symptom time series alone or combined with historical data.

| Symptom Only |  |  |  | Symptom + Historical |  |  |
| --- | --- | --- | --- | --- | --- | --- |
| Symptom | Optimal Window | Validation (rank) | Test | Optimal Window | Validation (rank) | Test |
| Ageusia | 4 | 91.2 (5.0) | 61.2 | 7 | 28.2 (9.0) | 16.6 |
| Anosmia | 9 | 83.2 (2.0) | 55.4 | 7 | 19.6 (7.0) | 12.2 |
| Chills | 4 | 91.7 (6.0) | 57.7 | 7 | 15.8 (6.0) | 11.4 |
| **Fever** | 3 | 115.0 (8.0) | 66.9 | **8** | **11.7 (1.0)** | **8.7** |
| Pneumonia | 3 | 130.6 (9.0) | 70.5 | 6 | 12.6 (3.0) | 11 |
| SOB | 3 | 114.6 (7.0) | 68.5 | 6 | 13.4 (4.0) | 8.3 |
| Dysgeusia | 4 | **78.6 (1.0)** | **59.8** | 7 | 25.4 (8.0) | 17 |
| Hypoxemia | 6 | 90.1 (4.0) | 62.6 | 8 | 15.6 (5.0) | 9.6 |
| Hypoxia | 6 | 84.8 (3.0) | 60.8 | 8 | 11.8 (2.0) | 9.6 |

In summary, our results suggested that developing SLSTM models trained using historical data and search trends for pneumonia yielded improvements in predicting 7 and 14 -day-ahead COVID-19 cases. Including search trends for fever in the development of the SLSTM models for forecasting 7-day-ahead COVID-19 deaths improved the performance. However, the model did not perform the best on the evaluation set. Thus, we failed to identify it as the optimal model. Finally, incorporating search trends for fever in training the SLSTM models led to improvement in the performance of the best model for predicting 14-day-ahead COVID-19 deaths. Next, we show that including search trends for two or three symptoms (instead of just one) yielded consistent improvements in the performance of the learned models. Besides, it also improved the agreement between the validation and test sets for identifying the best performing models.

### Further Improved forecasting of COVID-19 using Google search trends for more than one COVID-19 symptom

Results reported in Tables 3-6 demonstrated the viability of including Google search trends for one COVID-19 related symptom in training the SLSTM models. Here, we assessed whether including Google search trends for two or three COVID-19 related symptoms could further improve the predictive performance of the models. In this experiment, for each prediction task, we ranked the symptoms based on their MAPE scores obtained using the validation set for the bivariate SLSTM models (See Tables 3-6). Then, we considered the combinations of historical data with the top two and top three symptoms data series. Table 7 shows the best performing SLSTM models trained using historical data and Google search trends for up to three symptoms. For 7 and 14 -day-ahead forecasting of COVID-19 transmission, the best performed models used a window of size equals 3 and utilized the historical data and the top three symptoms data series to achieve the best reported MAPE values of 3.2% and 5.6%, respectively. Although including the time series for the top three symptoms relevant for forecasting COVID-19 mortality provided SLSTM models with better performance than those trained using historical data only, the best performing models for 7 and 14 -day-ahead forecasting of COVID-19 mortality were obtained using one and two symptoms, respectively. Interestingly, all optimal models highlighted in bold in Table 7 had their lowest MAPE values on both validation and test sets. Another interesting observation is that results in Table 7 suggest that forecasting COVID-19 mortality is more challenging than forecasting COVID-19 transmission: training COVID-19 mortality forecasting models required longer window sizes that span 6 or 7 days and MAPE scores for these models is worse than those for the COVID-19 transmission forecasting models.

**Table 7:** Performance of the best SLSTM models trained using historical data plus Google search trends for 0-3 COVID-19 related symptoms.

| **Task** | **Symptoms** | **Optimal Window** | **Validation** | **Test** |
| --- | --- | --- | --- | --- |
| Cases-7 | None | 9 | 7.4 | 6.6 |
|  | Pneumonia | 3 | 5.2 | 3.8 |
|  | Pneumonia + SOB | 3 | 5.3 | 3.2 |
|  | **Pneumonia + SOB + Fever** | **3** | **5.18** | **3.2** |
| Cases-14 | None | 9 | 11.8 | 8.8 |
|  | Pneumonia | 5 | 9.6 | 7.4 |
|  | Pneumonia + Fever | 3 | 9.8 | 6.5 |
|  | **Pneumonia + Fever + SOB** | **3** | **9.5** | **5.6** |
| Mortality-7 | None | 6 | 7.3 | 4.8 |
|  | **Hypoxia** | **6** | **6.7** | **4.7** |
|  | Hypoxia + Chills | 5 | 8 | 6.1 |
|  | Hypoxia + Chills + SOB | 4 | 8.2 | 5.2 |
| Mortality-14 | None | 7 | 13.1 | 11.4 |
|  | Fever | 8 | 11.7 | 8.7 |
|  | **Fever + Hypoxia** | **7** | **11.4** | **7.8** |
|  | Fever + Hypoxia + Pneumonia | 7 | 11.6 | 8.6 |

### Analysis of state-level predictions of the best performing models

State-level predictions of the four best performing models highlighted in Table 7 are provided in Supplementary files 1-4 and Tables S1-S4. Fig. 4 shows sample test results for three states (AZ, UT, and CA) on predicting 7-day-ahead COVID-19 confirmed cases, where true and predicted trajectories are highlighted in blue and red, respectively. Based on this figure, we categorized the 51 states into one of three categories based on the relationship between true and predicted trajectories: i) over-estimated group, where at least 90% of the predictions are over-estimated; ii) under-estimated group, where at least 90% of the predictions are under-estimated; and iii) others. For the four classification tasks considered in this study, Fig. 5 shows the categorization of states into three groups such that states with over-estimated predictions are highlighted in red, states with under-estimated predictions are highlighted in yellow and the remaining states are highlighted in gray. Interestingly, the US states with shared borders were more likely to be assigned to the same category. An interesting exception is WA, which always appeared as an isolated state belonging to the over-estimated category.

**Figure 4.**
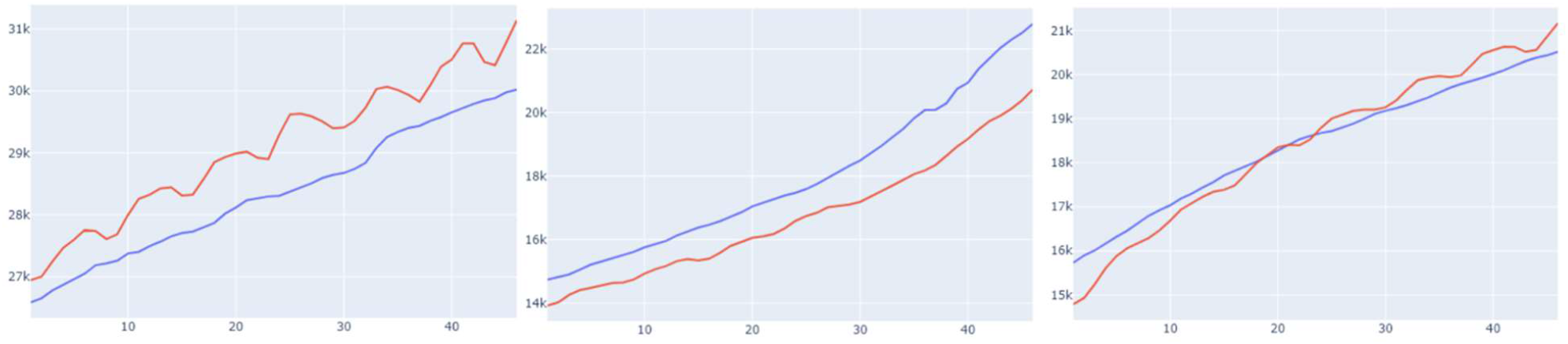
Examples of (left) over-estimated predictions (middle) under-estimated predictions (right) other predictions.

**Figure 5.**
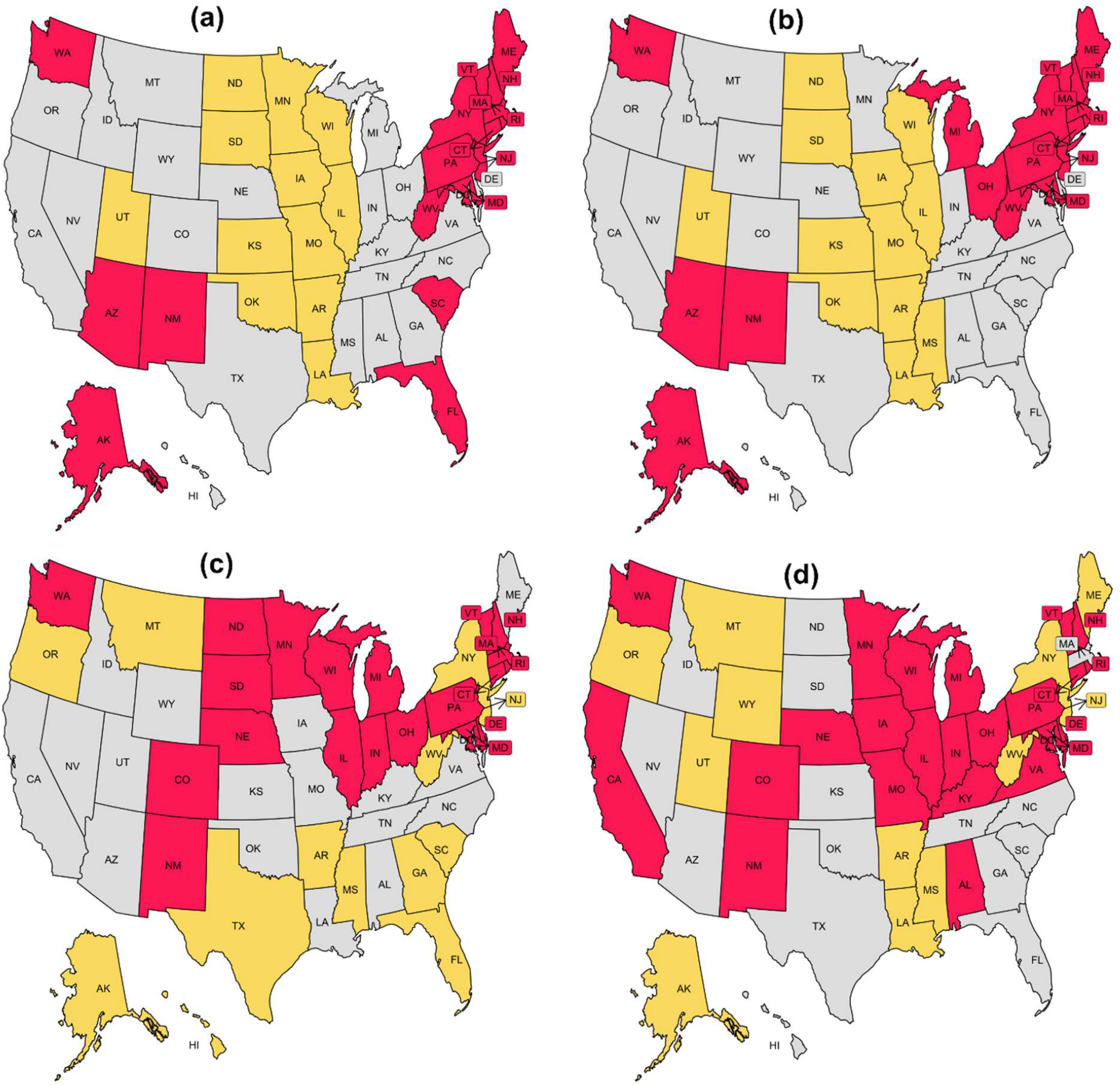
US States with over-estimated (red), under-estimated (yellow), and others (gray) for the four prediction tasks: (a) Cases-7; (b) Cases-14; (c) Mortality-7; (d) Mortality-14.

To get insights into when we should expect our best performing models to have over/under -estimated predictions, we examined the trajectories of the member states in the three groups for each of the four tasks (See Supplementary Figs. S1-S4). These four figures are also summarized in Fig. 6 by representing the trajectories in each cluster using their pointwise mean curve. For COVID-19 cases prediction tasks, Cases-7 and Cases-14, we found that, during the test interval, states in the over-estimated groups had mean curves at the top of the entire data mean curve, while states in the ‘others’ group had a mean curve that is close to the whole data mean curve. Surprisingly, this observation did not apply to the COVID-19 death prediction tasks, Mortality-7 and Mortality-14.

**Figure 6.**
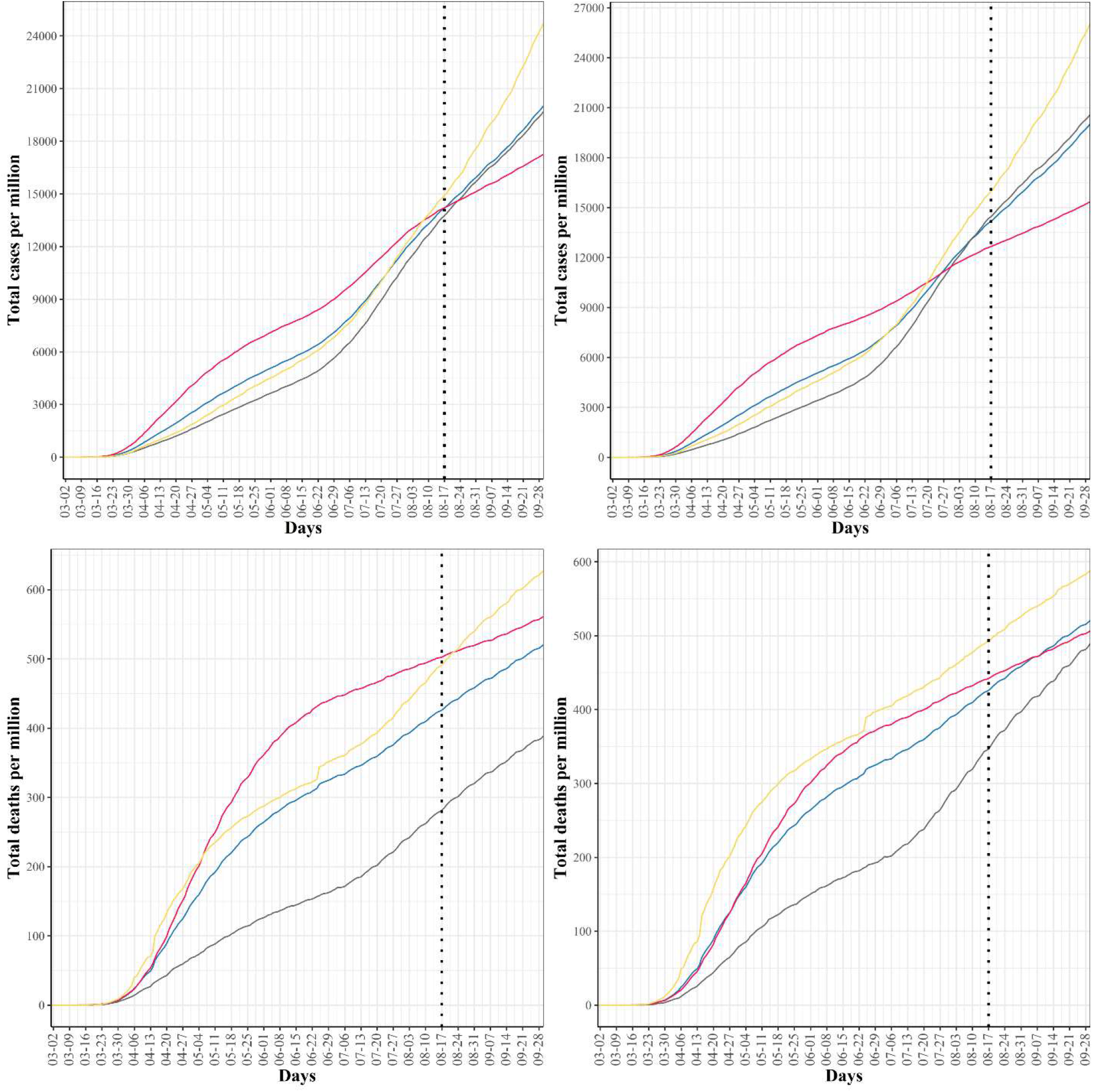
Mean curve for: over-estimated (red); under-estimated (yellow); others (gray); and all (blue) US states. Vertical dotted line indicates the beginning of the test time series.

## Discussion

Recent advances in machine learning based time series forecasting, particularly deep LSTM networks, enabled the development of complex non-linear deep architectures for modeling dynamics and long-term dependencies in real-world time series data. To date, several COVID-19 forecasting models have been developed using several variants of the LSTM architecture (e.g. [21, 49-51]). However, the size of the test data in existing studies was relatively small because it equals the number of days in the test time interval. Fortunately, this is not the case in this study. Though our test data spans 45 days interval (from August 17^th^ to end of September), the size of our test data was 51 *x* 45 = 2295 samples because we experimented with the US data at the state level. Generally, a large test data is required to accurately evaluate the performance of a prediction model [52]. Thus, or performance estimates are thought to be more robust as the size of our test data is more than 10 times the size of the test data used in related studies.

Since the beginning of the COVID-19 pandemic in 2020, tracking and modeling its spread have gained considerable attention from health agencies. Therefore, it is not surprising that numerous computational methods for forecasting the transmission, mortality, and recovery of COVID-19 have been proposed in the literature. Among these methods, few methods incorporated other sources of relevant data such as Google trends [53, 54], climate [55], and mobility [56] in their forecasting systems. To the best of our knowledge, our proposed models are the first models that use queries from the Google COVID-19 symptoms database [43] to improve their predictive performance on forecasting COVID-19 transmission and mortality.

It is worse noting that our results demonstrated the added value of using Google search trends for COVID-19 related symptoms (i.e., fever, pneumonia, shortness of breath, and hypoxia) combined with historical data in forecasting COVID-19 confirmed and death cases. However, despite their strong and significant correlations with COVID-19 spread and death trajectories reported in [41], we found that SLSTM models trained using any of these symptoms alone yielded models with poor performance. In addition to the improvement in predictive performance obtained via using Google symptoms time series along with the historical data in training our models, another significant gain is improving the generalizability of the models on both validation and test sets (i.e., the best performing model on the validation set is the model with the best performance on the test set).

Our analysis of state-level predictions (Tables S1-S4) demonstrated high variability in the observed performance across various US states. This observation suggests that the identified Google search trends, though leading to the optimal overall performance, might not be the best for some US states. To overcome this limitation, we are interested in developing state-specific models, which is the subject of our ongoing work.

## Conclusions

We have developed deep learning models for forecasting COVID-19 infection and mortality in the US using historical data and Google search trends for COVID-19 related symptoms. Out of 422 symptoms included in the Google COVID-19 symptoms database [43], we have focused on the nine symptoms identified in [41] using dynamic correlation analysis. We then re-ranked these nine symptoms based on the performance of deep learning models trained using historical data and every single symptom. Finally, we used the top three symptoms to develop our final and best performing models. Our results suggest that Google search trends for the symptoms related to a target infectious disease could potentially improve the performance of the forecasting models for that disease. Our future work aims at: including more relevant time series data (e.g., mobility and climate) and assessing the improvement in performance for forecasting over longer intervals (e.g., 21 and 30 days); experimenting with other deep learning models for time series (e.g., Convolutional LSTM [57] and Bidirectional LSTM [47]); and developing state-specific forecasting models.

## Data Availability

State-level cumulative numbers of daily COVID-19 confirmed cases and deaths were obtained from the publicly available repository maintained by the Center for Systems Science and Engineering (CSSE) at Johns Hopkins University (JHU). The Google COVID-19 Search Trends Symptoms dataset is publicly available at https://github.com/google-research/open-covid-19-data/. The 2019 US Census data are available at https://www.census.gov/data.html.

## Supplementary Information

**Supplementary file 1**: Per-state predictions of the best performing model versus COVID-19 cases for the CASES-7 task.

**Supplementary file 2**: Per-state predictions of the best performing model versus COVID-19 cases for the CASES-14 task.

**Supplementary file 3**: Per-state predictions of the best performing model versus COVID-19 death cases for the Mortality-7 task.

**Supplementary file 4**: Per-state predictions of the best performing model versus COVID-19 death cases for the Mortality-14 task.

**Supplementary file 5**: Tables S1-S4

**Supplementary file 6**: Figures S1-S4

