## Supplementary figures and images for "Improved Prediction of COVID-19 Transmission and Mortality Using Google Search Trends for Symptoms in the United States"

### AK_prediction.png

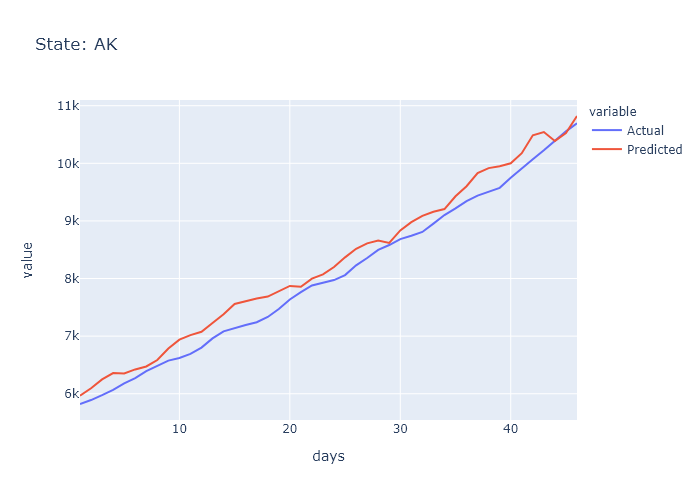

### AK_prediction.png

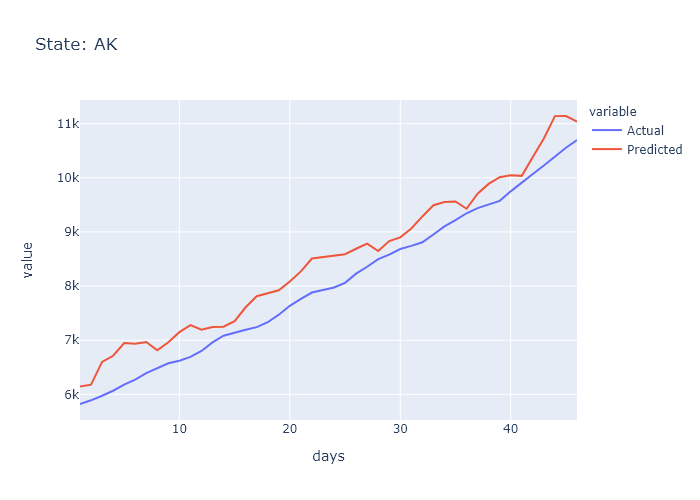

### AL_prediction.png

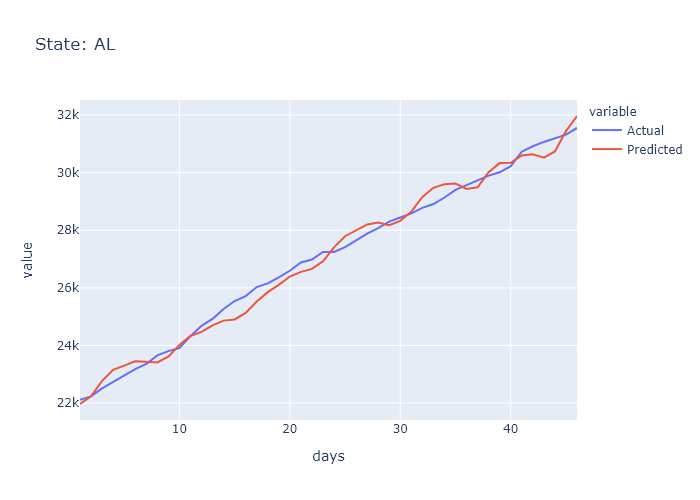

### AL_prediction.png

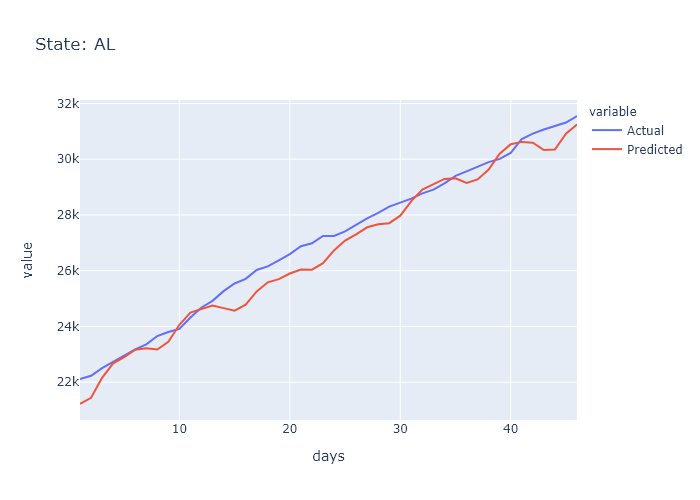

### AR_prediction.png

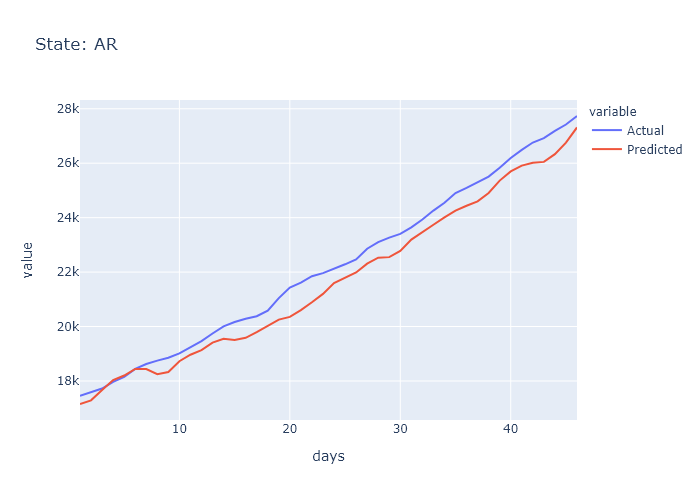

### AR_prediction.png

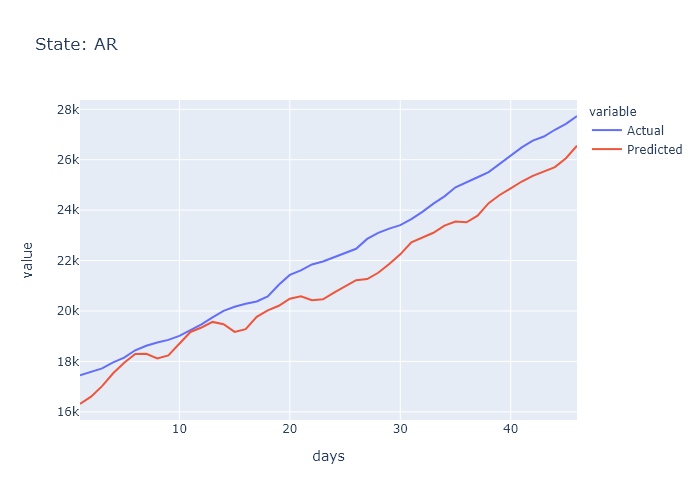

### AZ_prediction.png

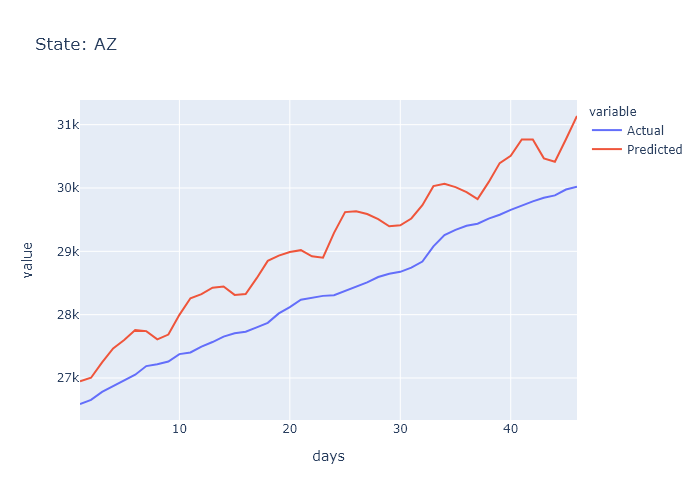

### AZ_prediction.png

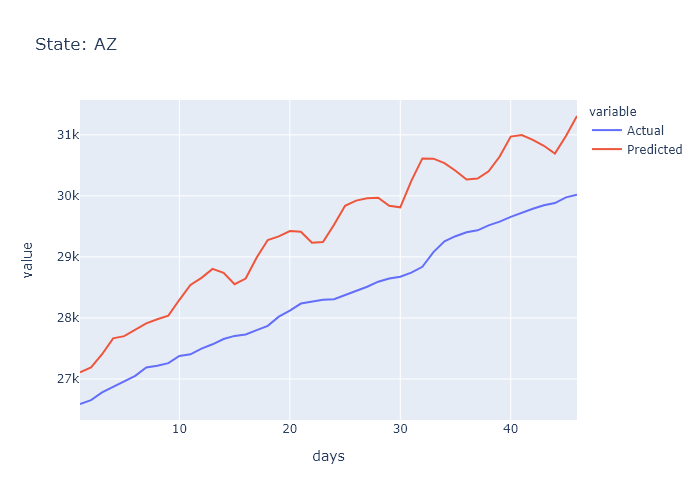

### CA_prediction.png

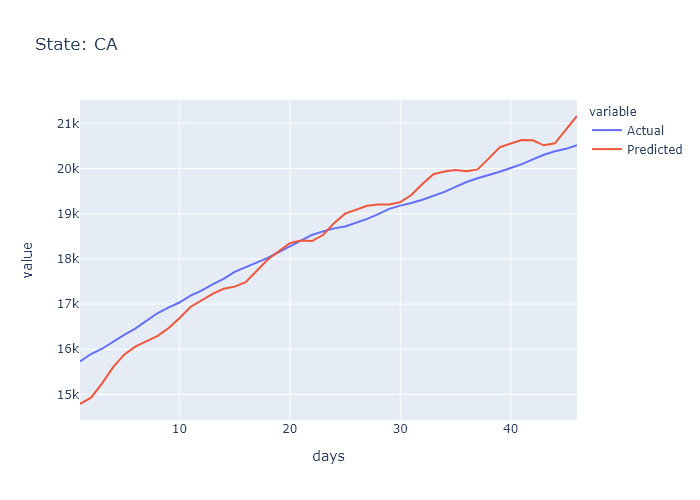

### CA_prediction.png

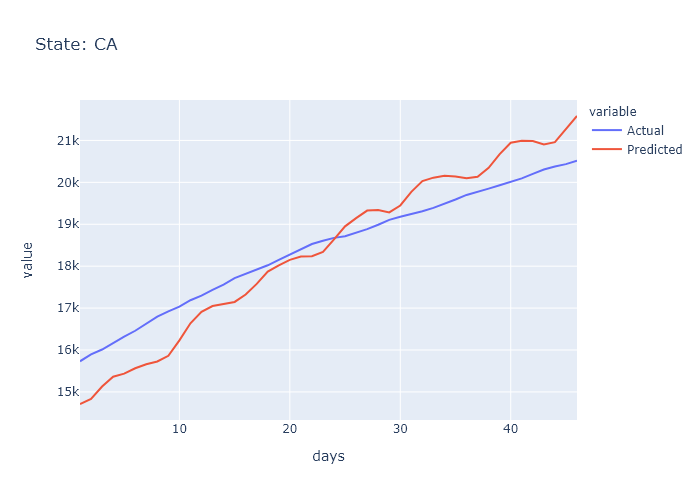

### CO_prediction.png

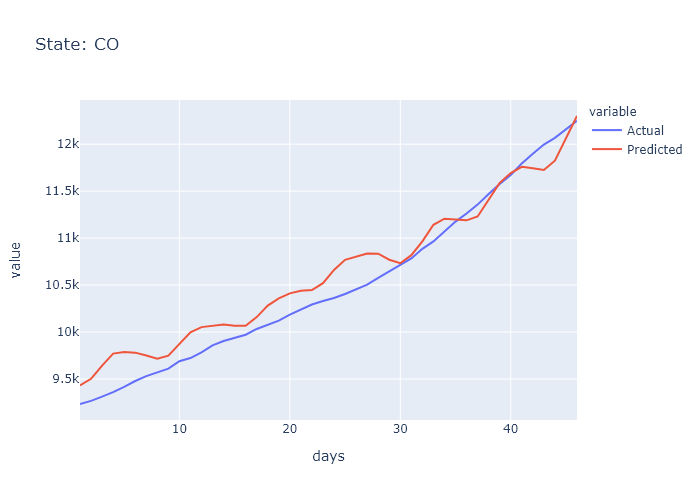

### CO_prediction.png

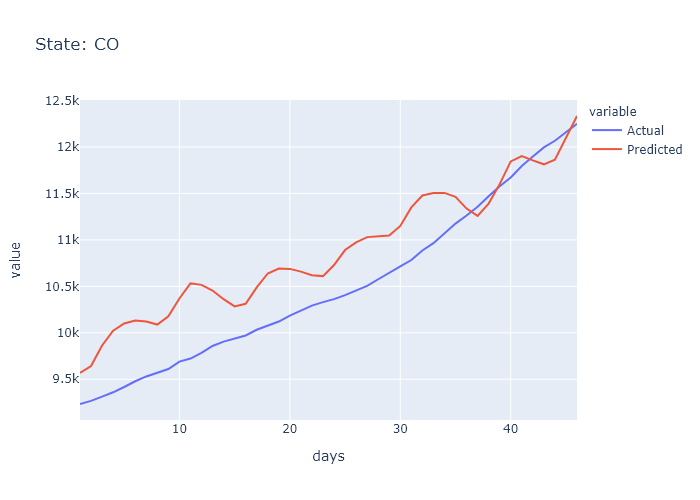

### CT_prediction.png

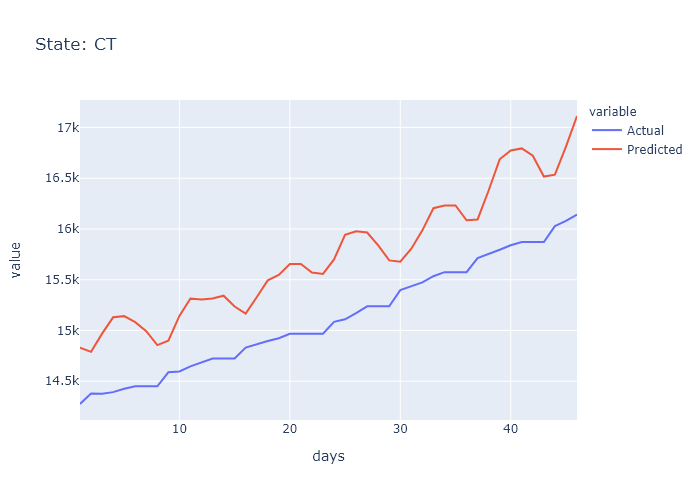

### CT_prediction.png

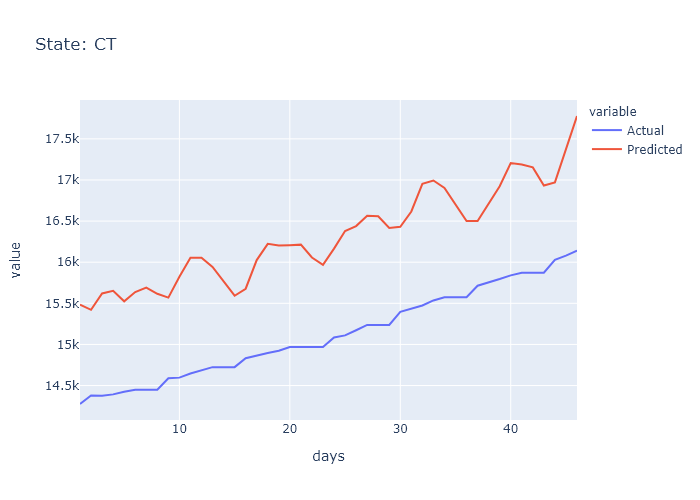

### DC_prediction.png

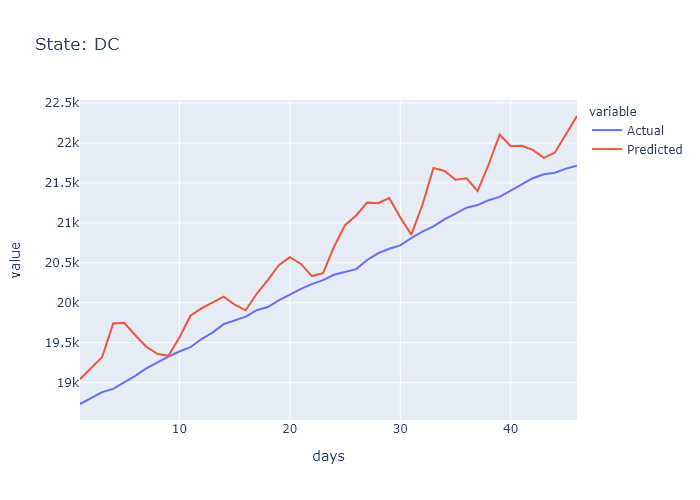

### DC_prediction.png

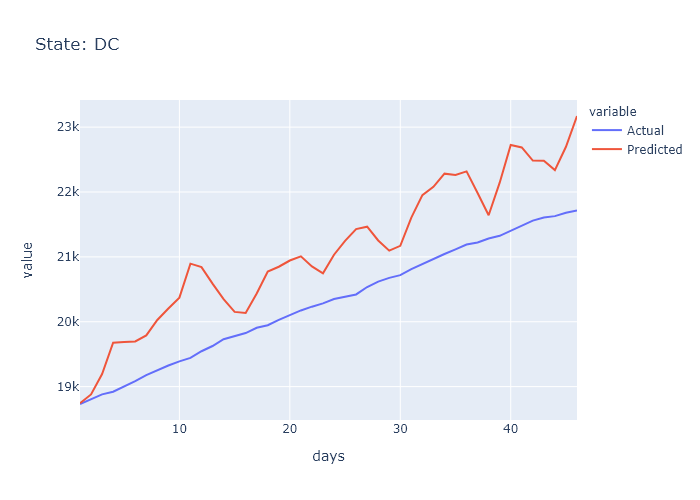

### DE_prediction.png

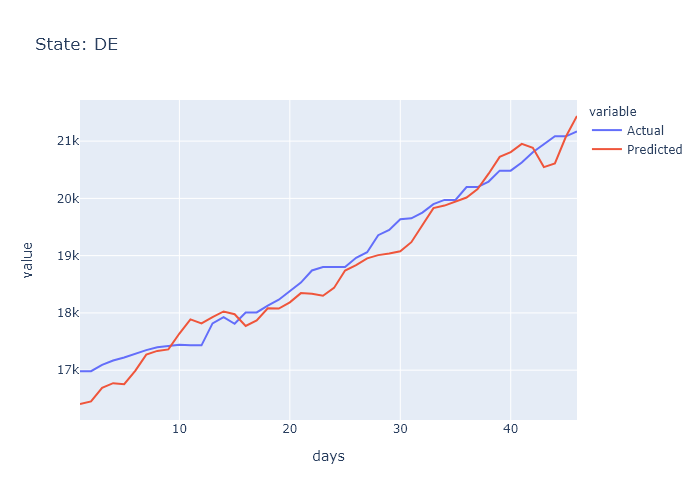

### DE_prediction.png

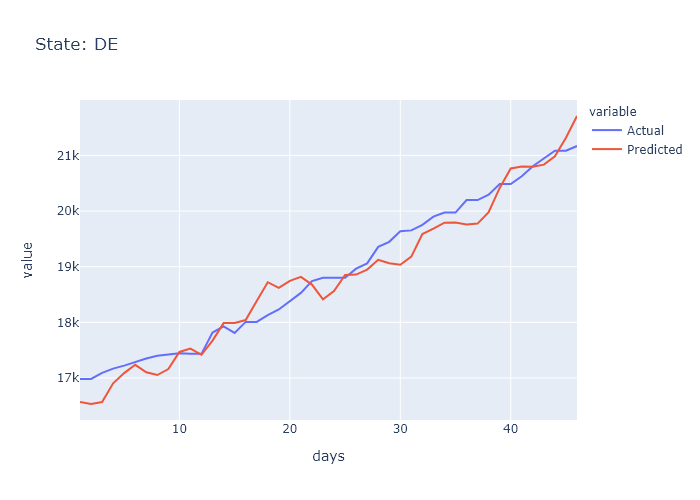

### FL_prediction.png

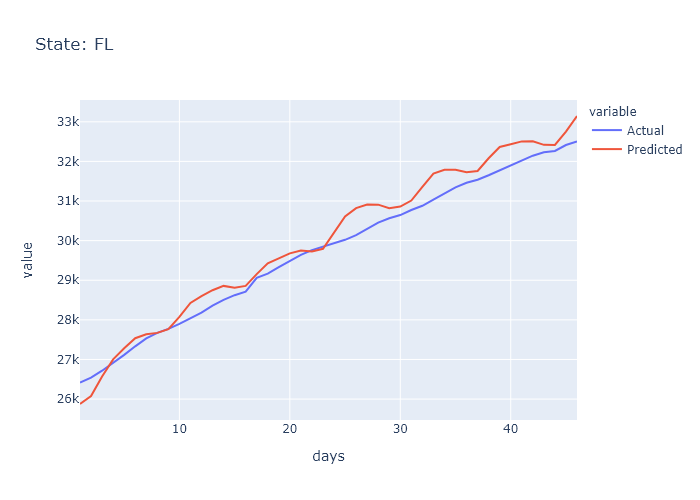

### FL_prediction.png

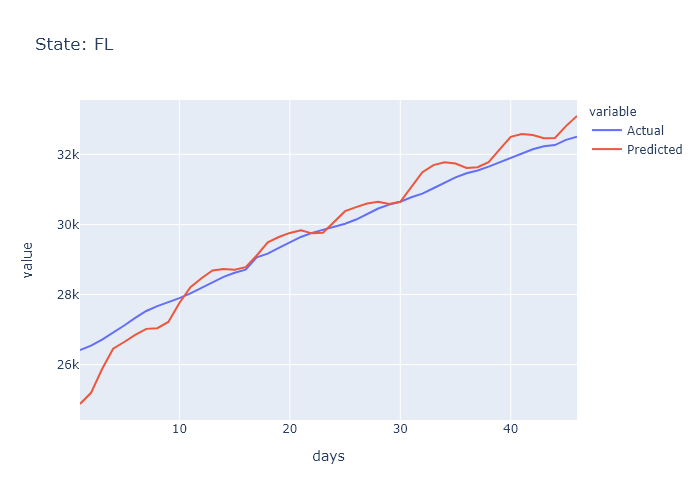

### GA_prediction.png

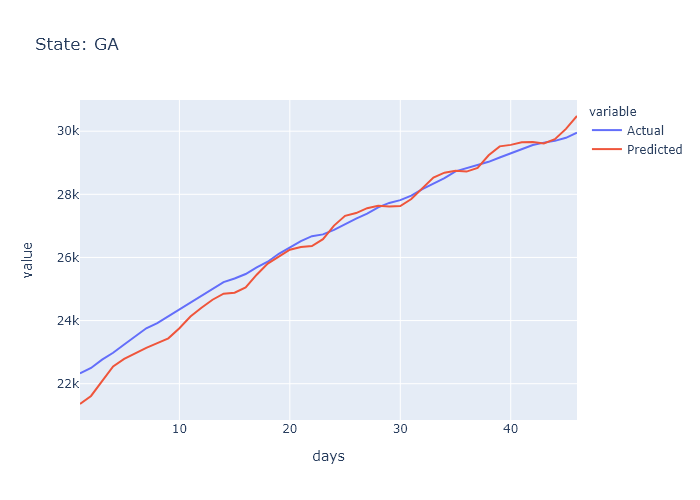

### GA_prediction.png

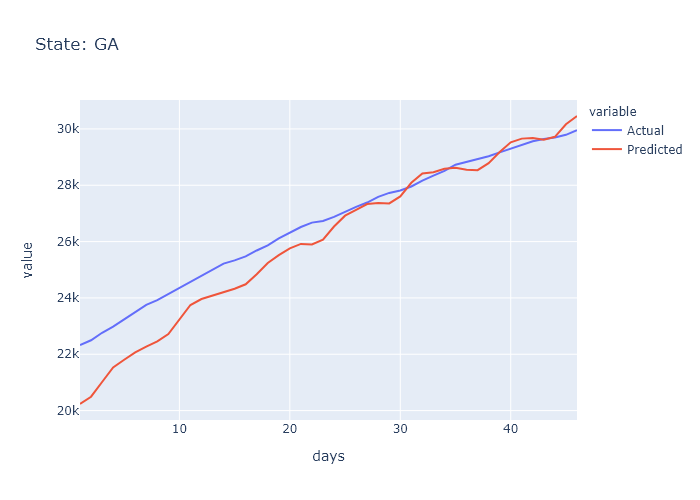

### HI_prediction.png

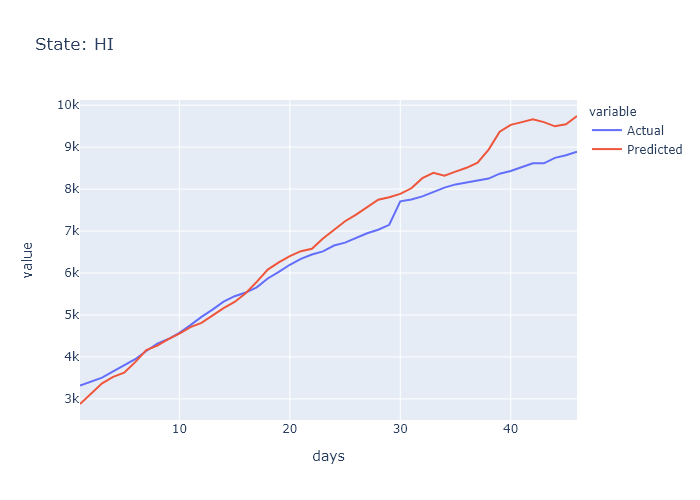

### HI_prediction.png

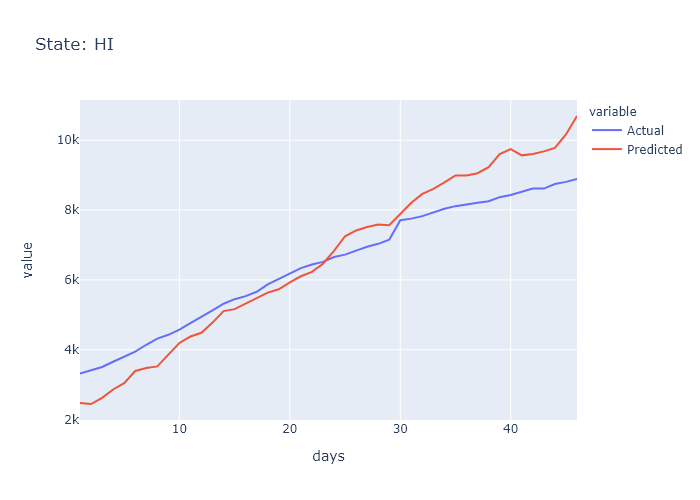

### IA_prediction.png

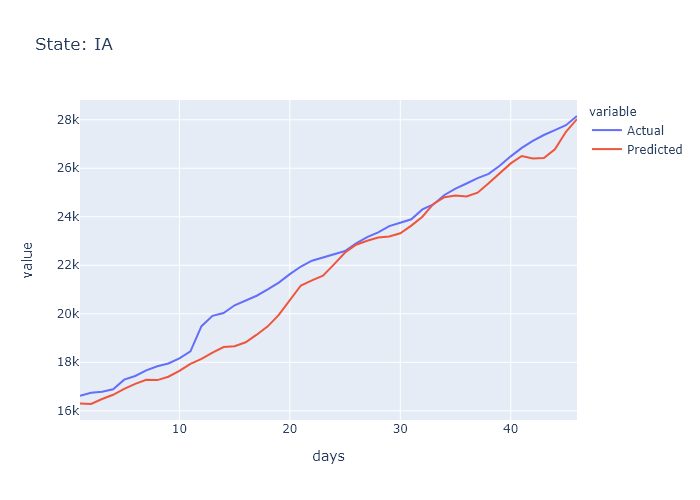

### IA_prediction.png

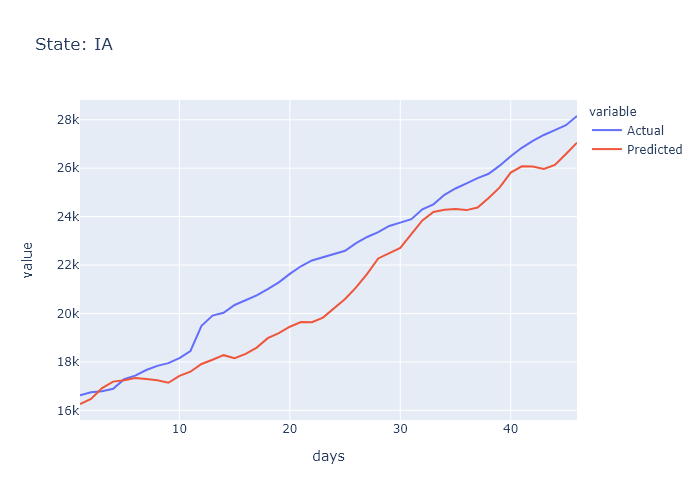

### ID_prediction.png

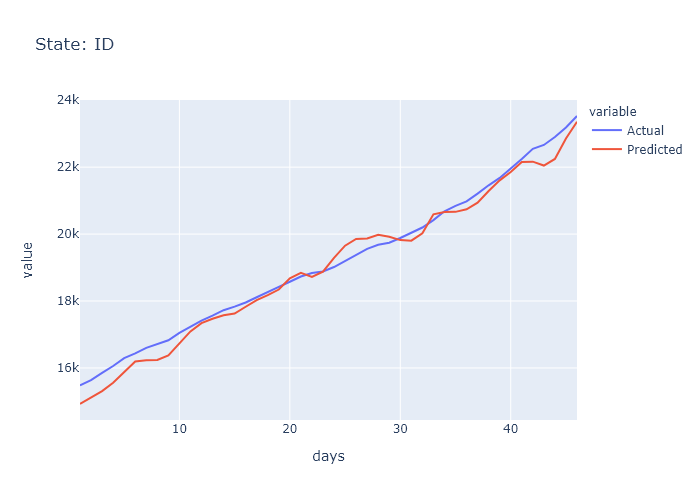

### ID_prediction.png

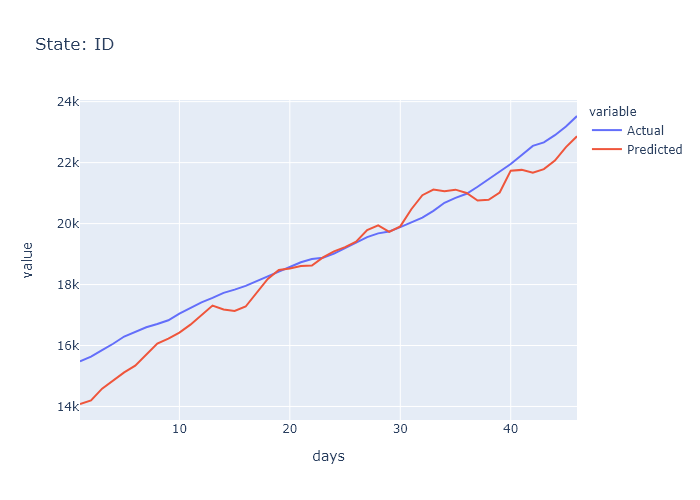

### IL_prediction.png

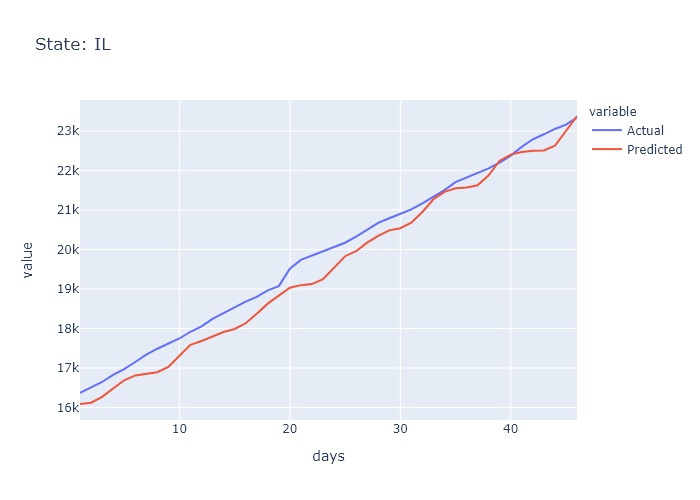

### IL_prediction.png

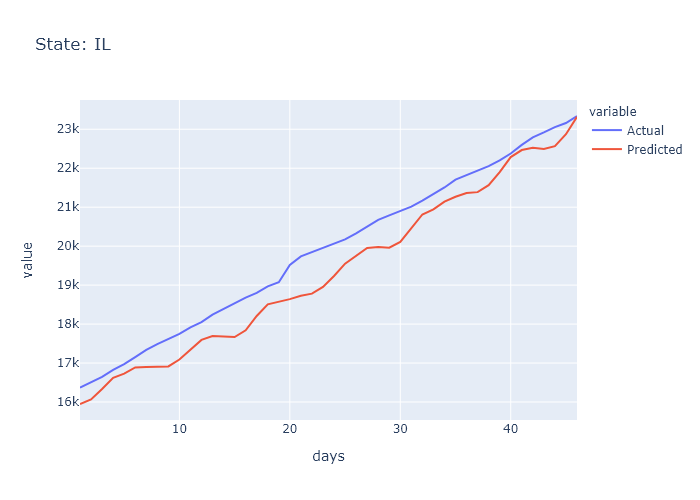

### IN_prediction.png

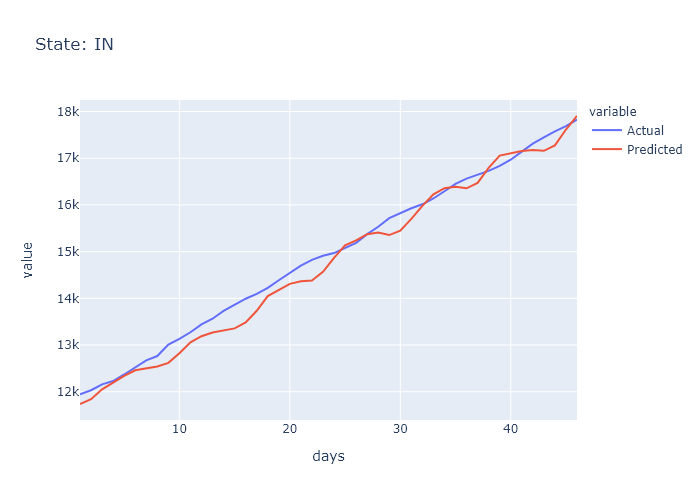

### IN_prediction.png

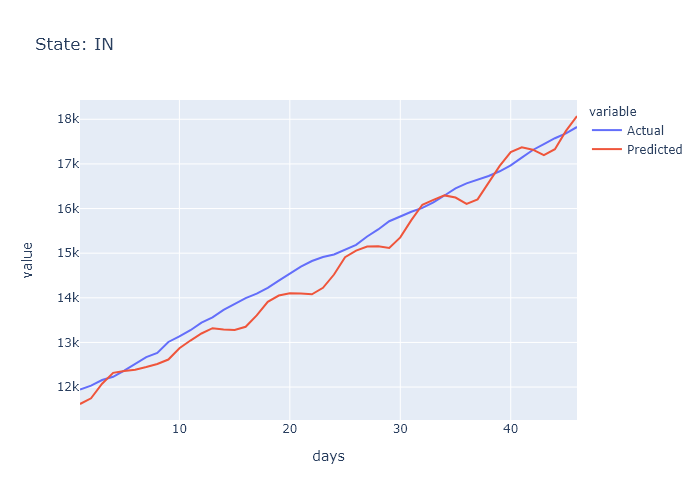

### KS_prediction.png

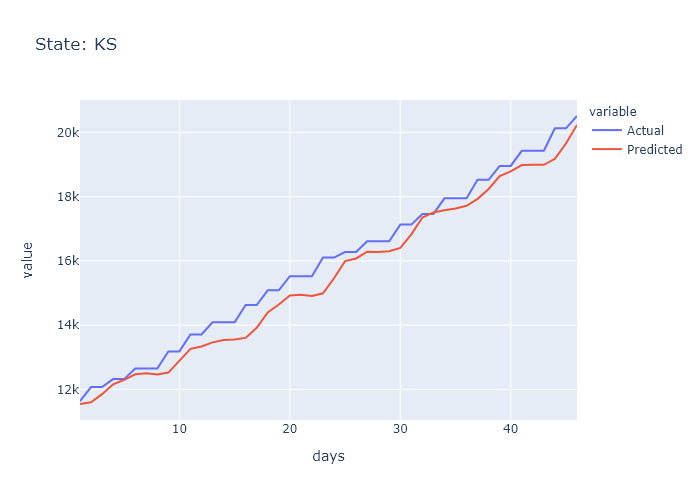

### KS_prediction.png

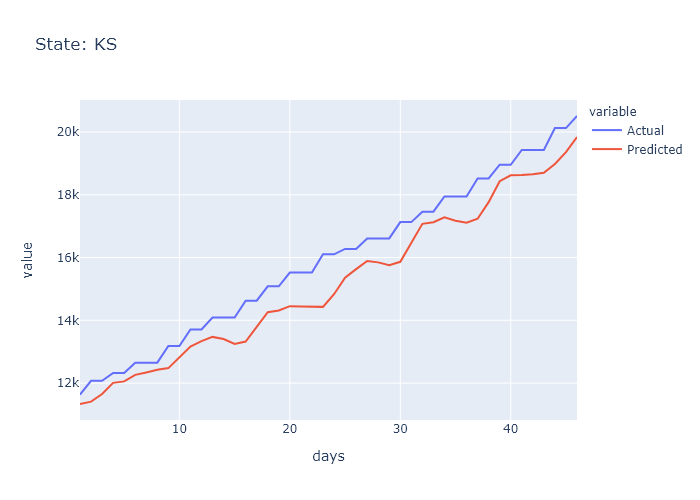

### KY_prediction.png

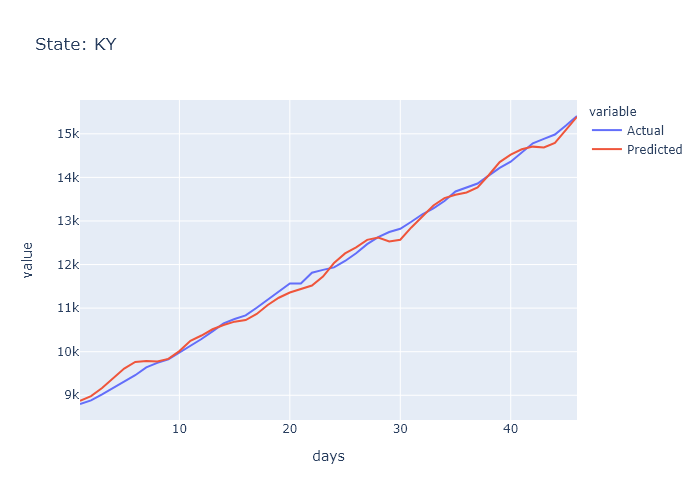

### KY_prediction.png

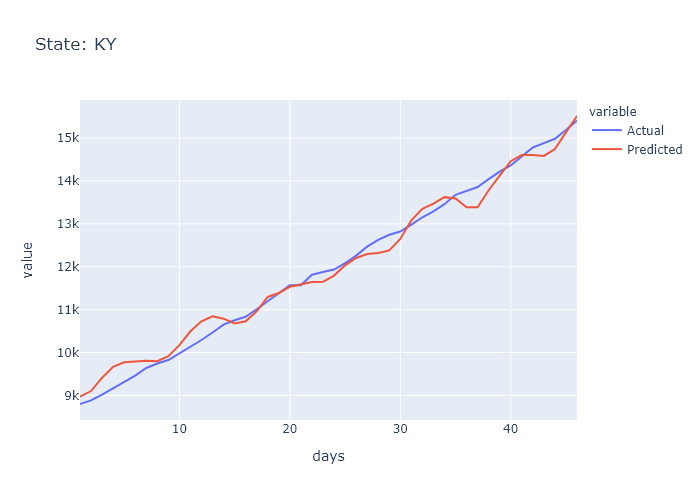

### LA_prediction.png

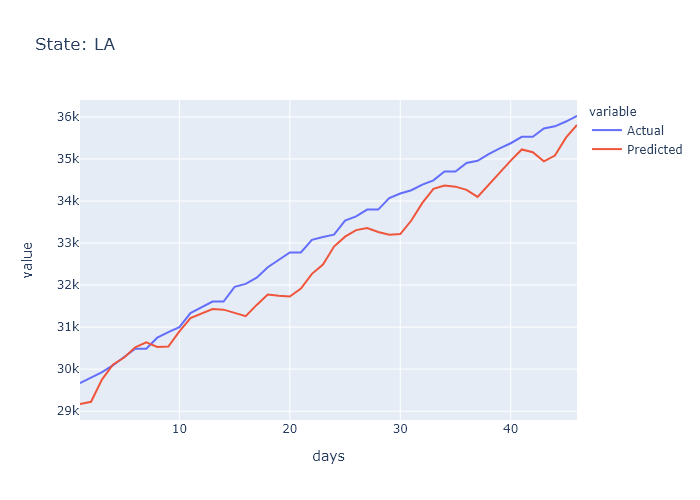

### LA_prediction.png

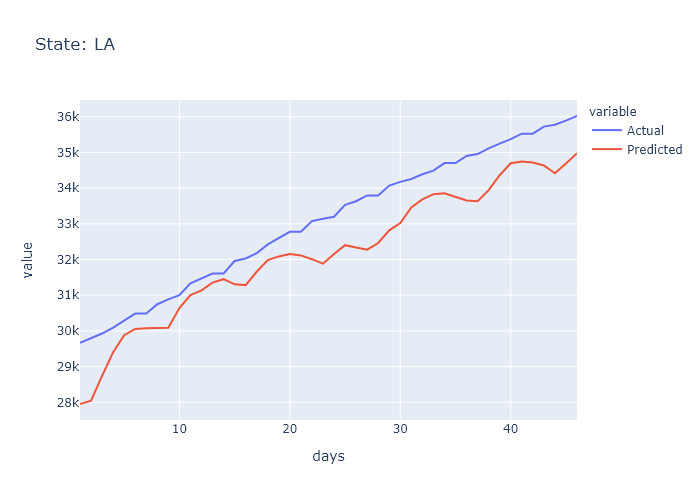

### MA_prediction.png

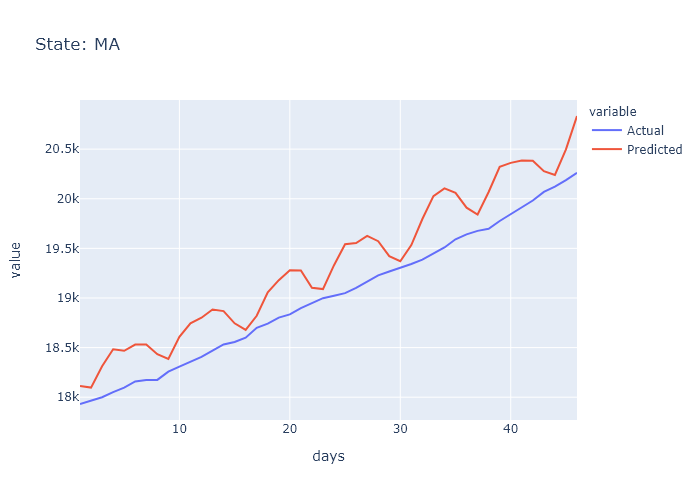

### MA_prediction.png

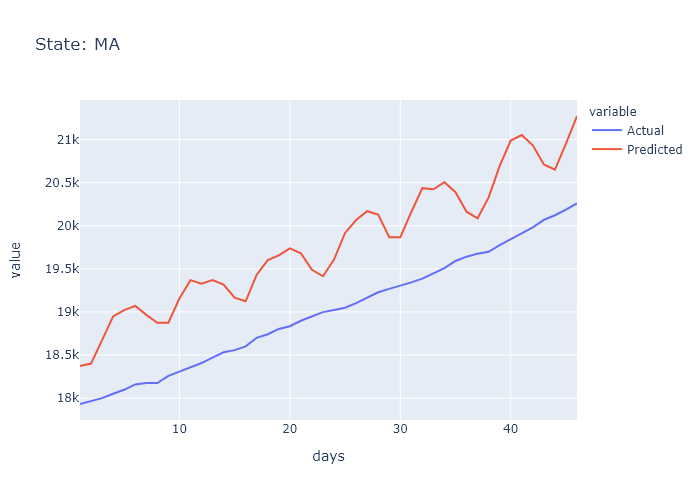

### MD_prediction.png

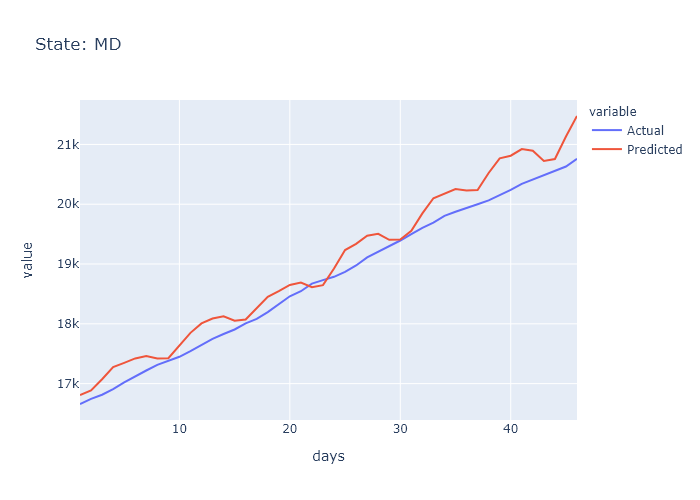

### MD_prediction.png

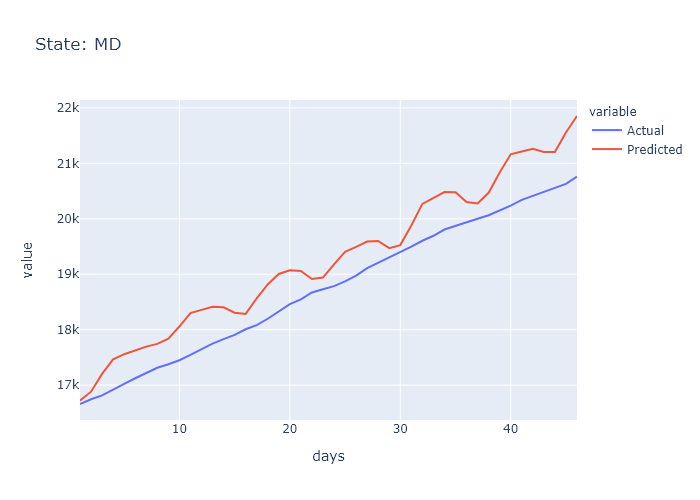

### ME_prediction.png

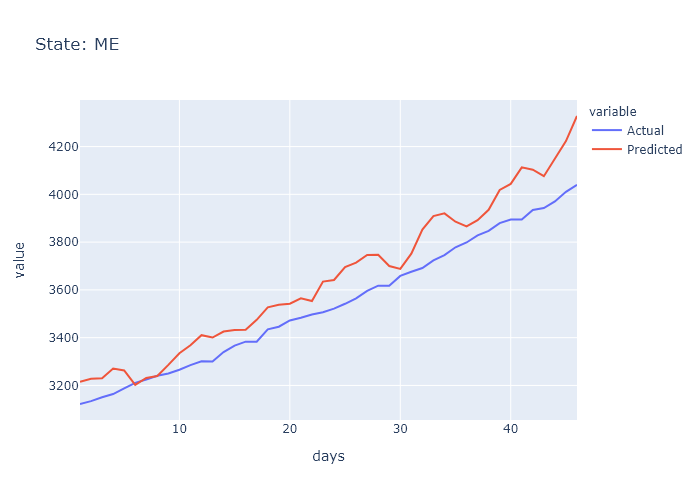

### ME_prediction.png

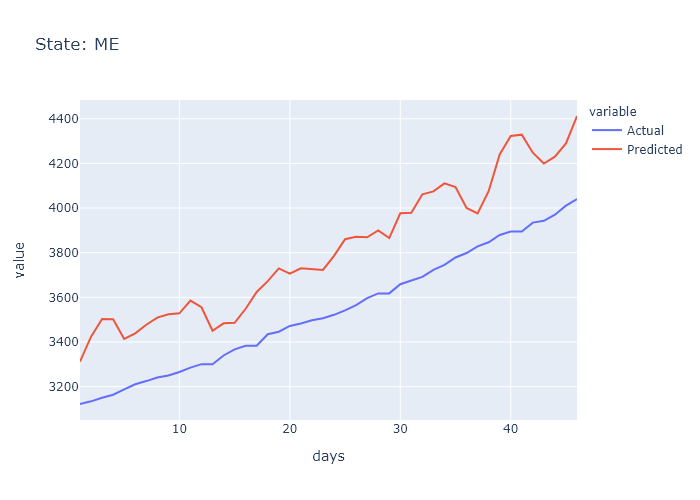

### MI_prediction.png

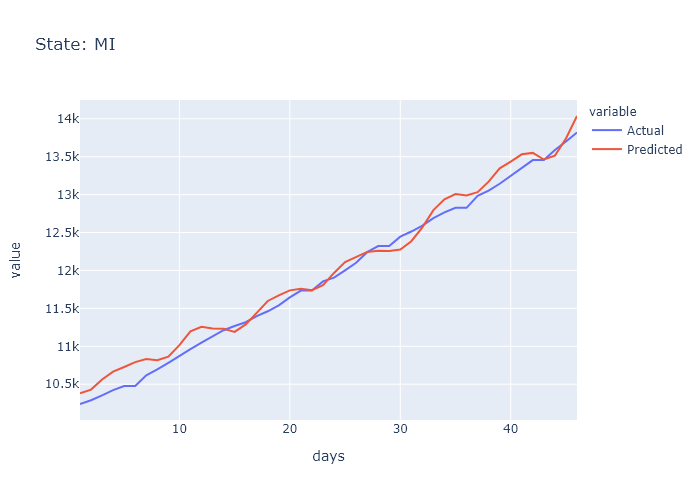

### MI_prediction.png

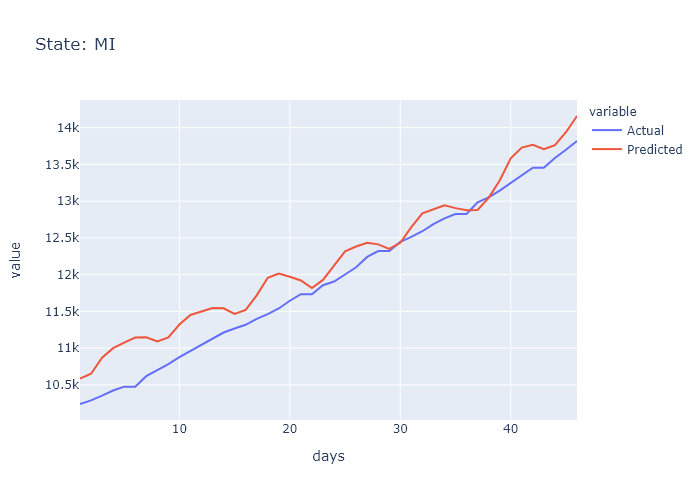

### MN_prediction.png

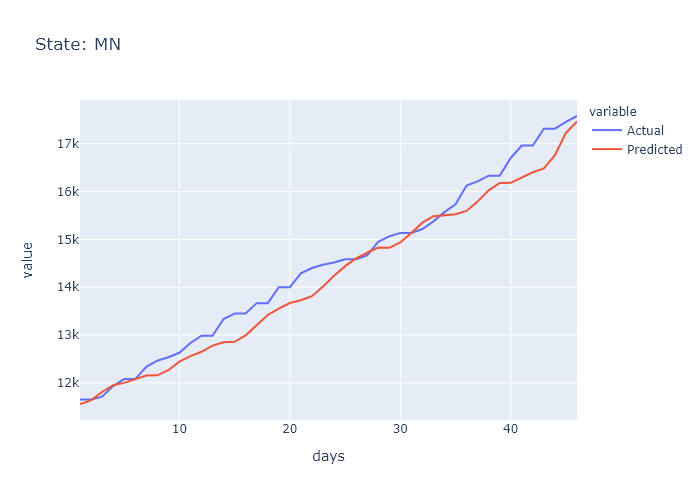

### MN_prediction.png

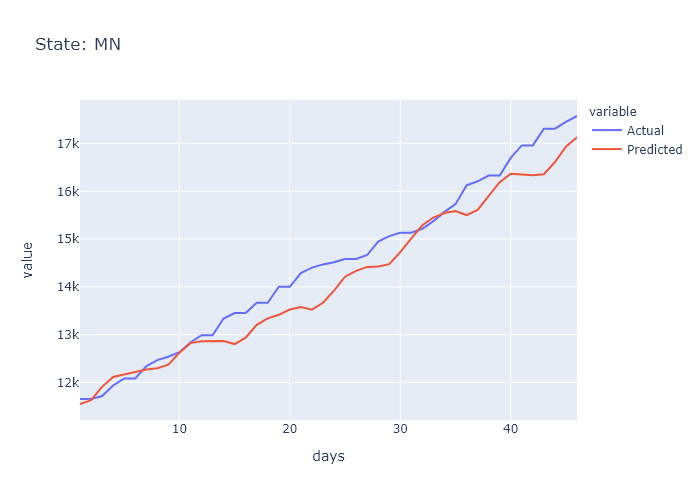

### MO_prediction.png

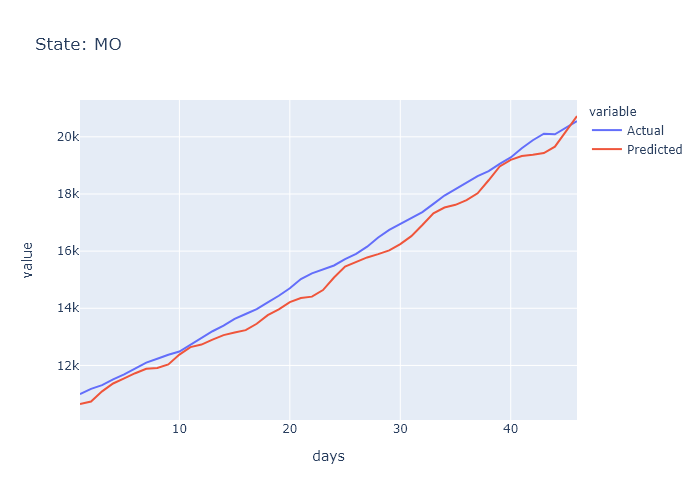

### MO_prediction.png

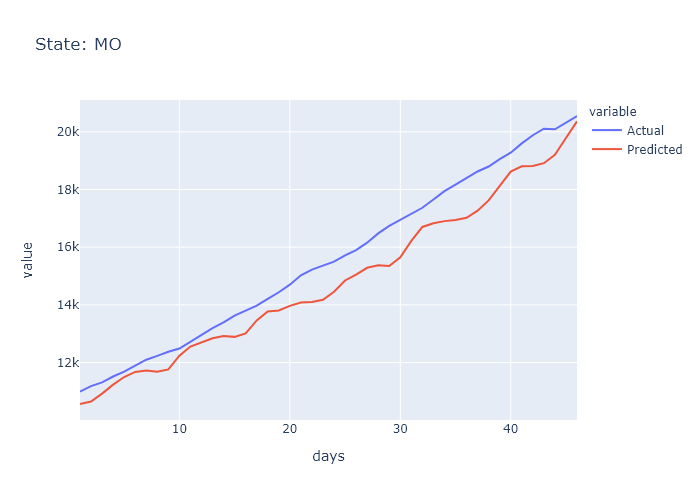

### MS_prediction.png

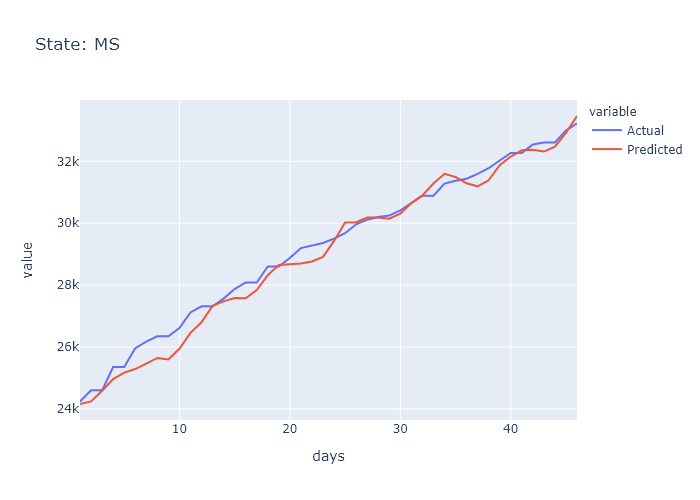

### MS_prediction.png

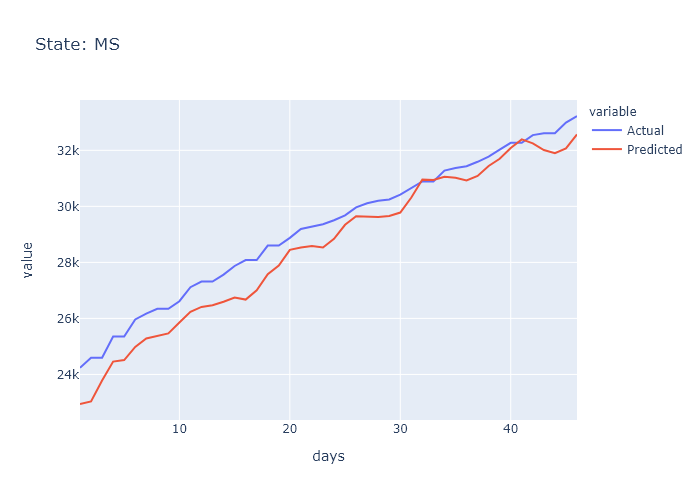

### MT_prediction.png

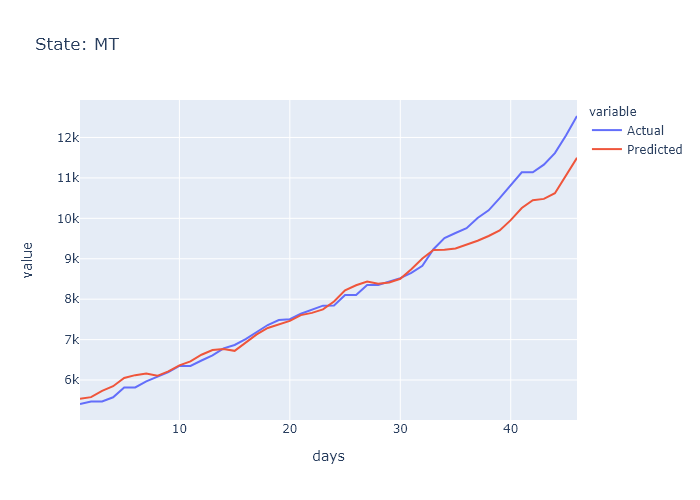

### MT_prediction.png

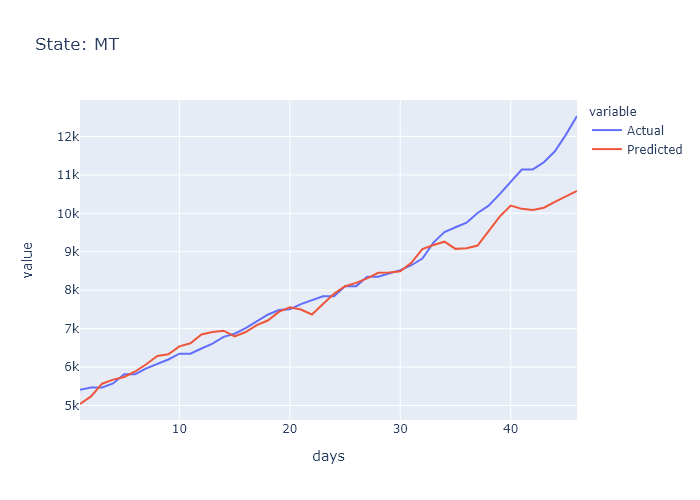

### NC_prediction.png

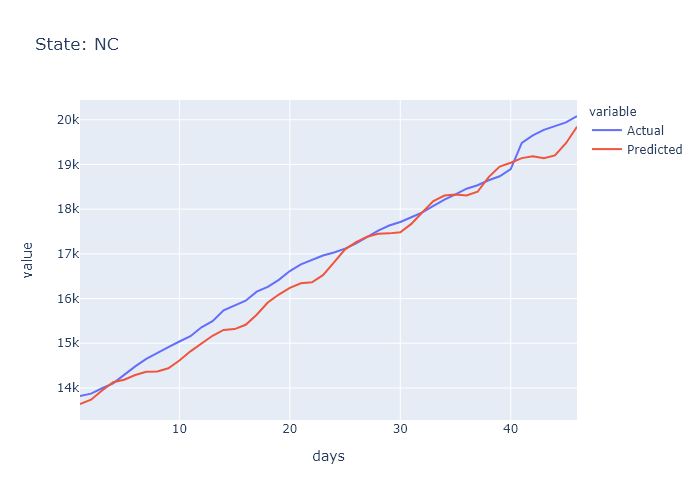

### NC_prediction.png

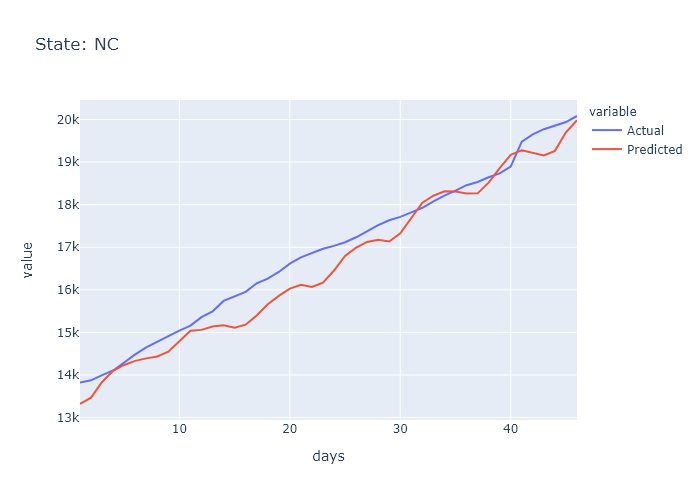

### ND_prediction.png

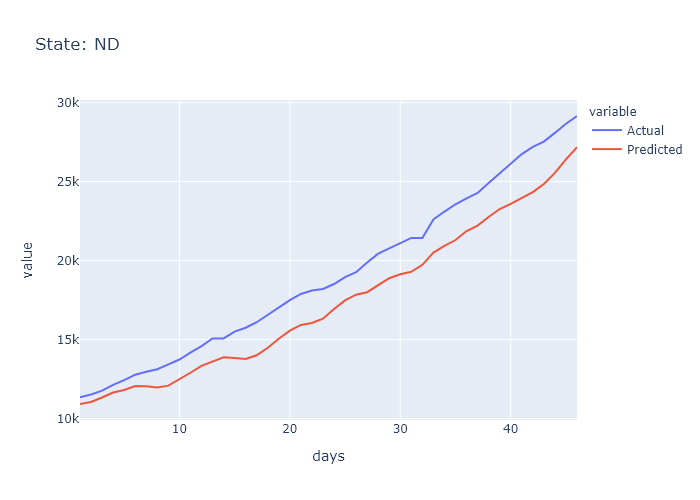

### ND_prediction.png

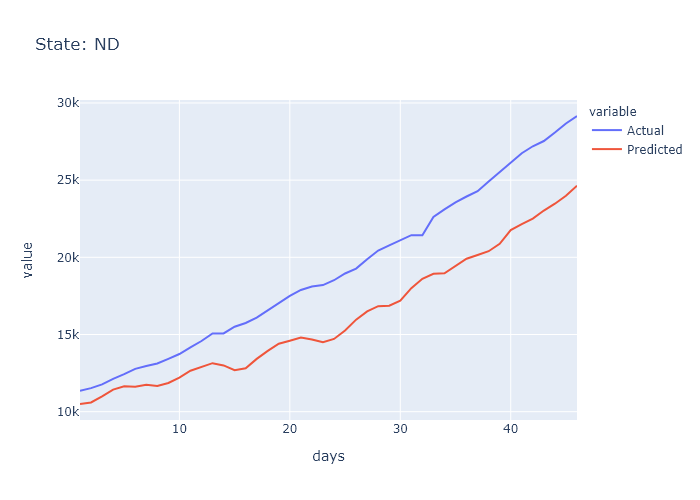

### NE_prediction.png

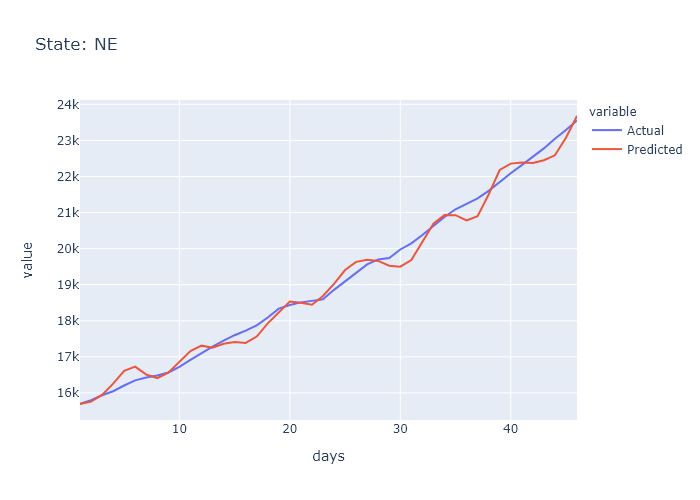

### NE_prediction.png

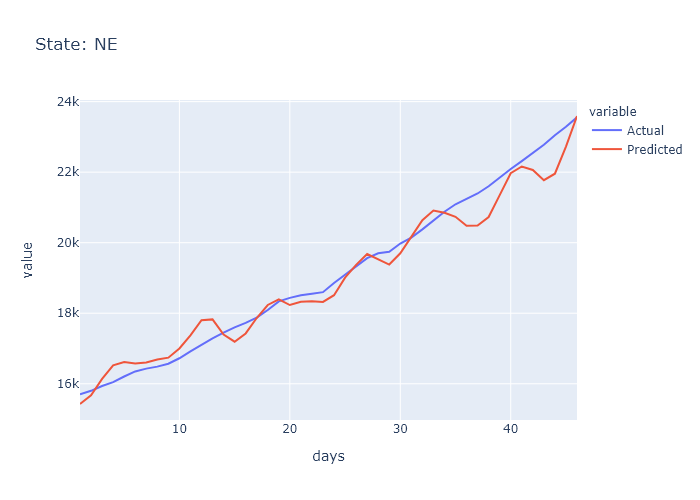

### NH_prediction.png

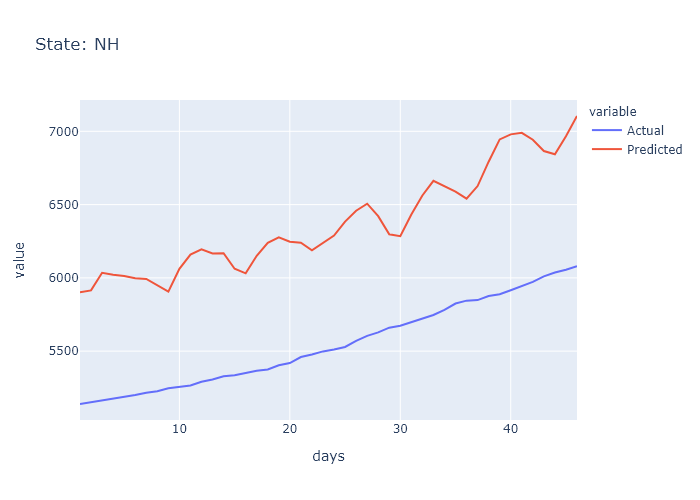

### NH_prediction.png

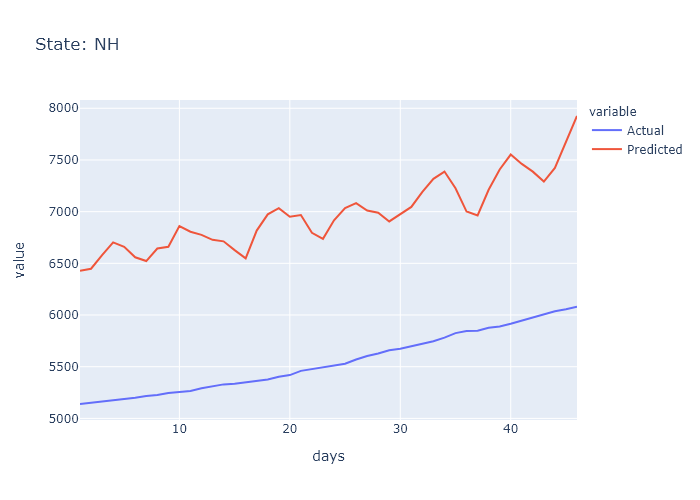

### NJ_prediction.png

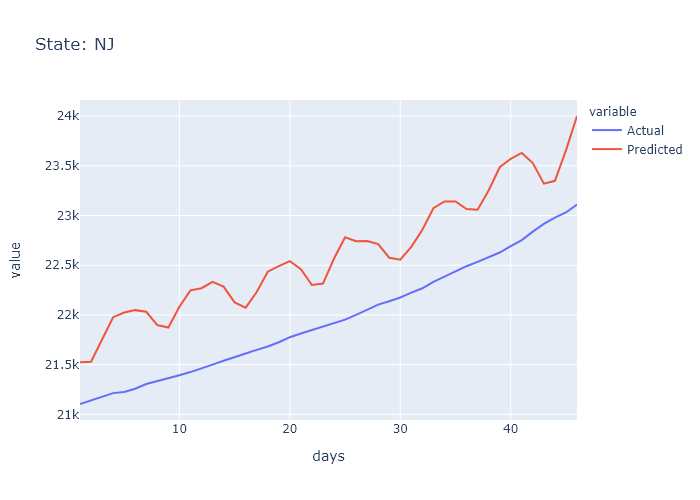

### NJ_prediction.png

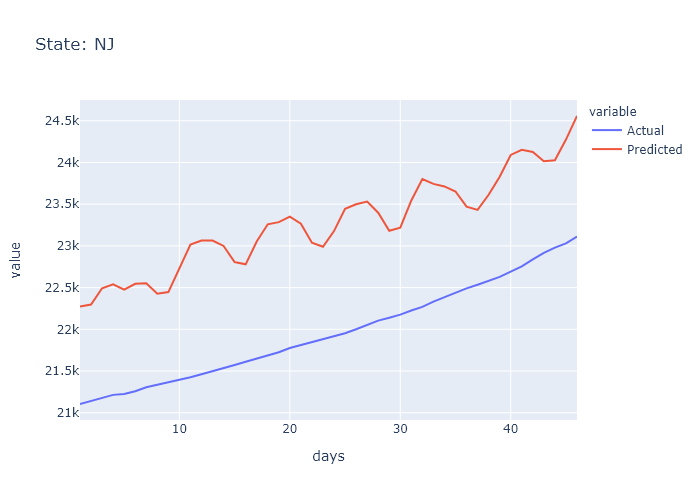

### NM_prediction.png

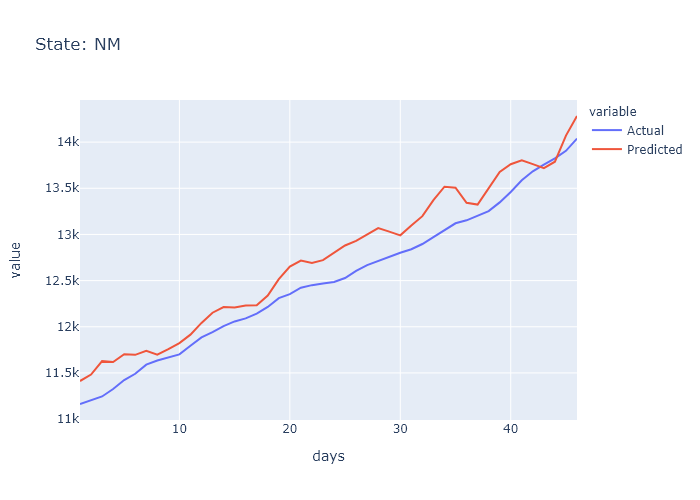

### NM_prediction.png

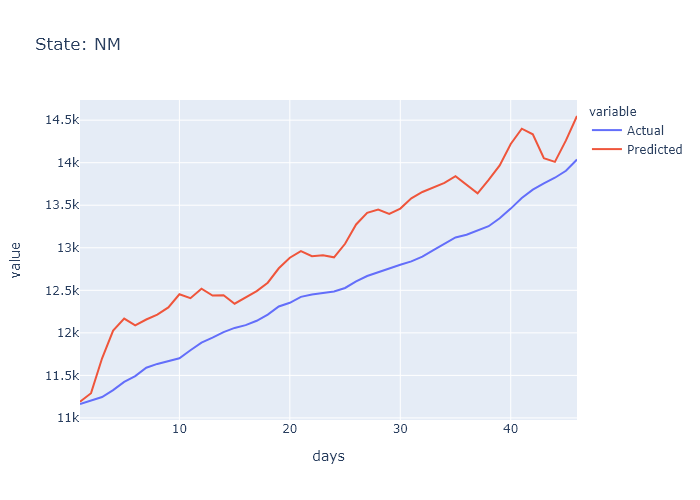

### NV_prediction.png

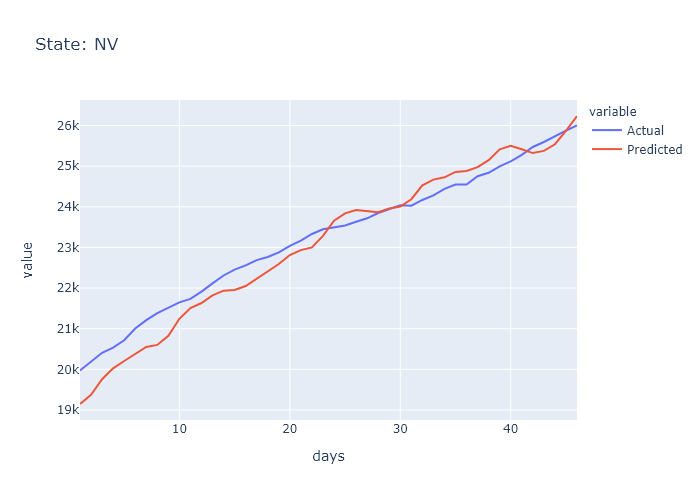

### NV_prediction.png

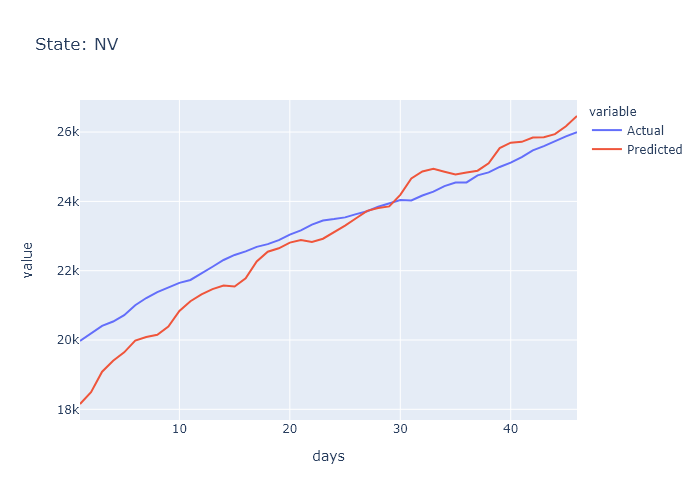

### NY_prediction.png

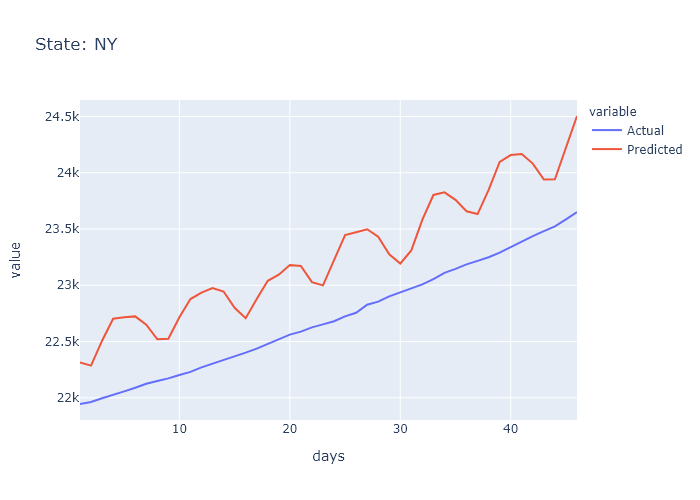

### NY_prediction.png

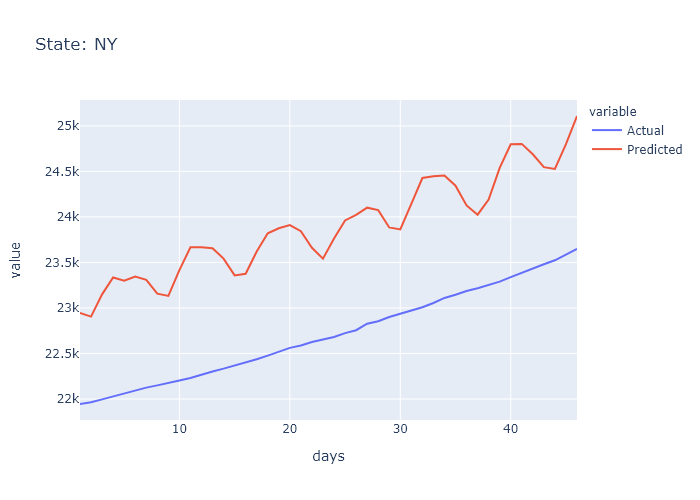

### OH_prediction.png

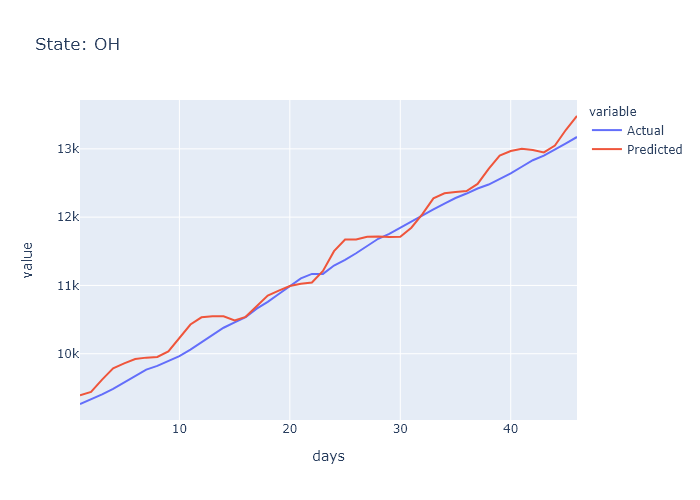

### OH_prediction.png

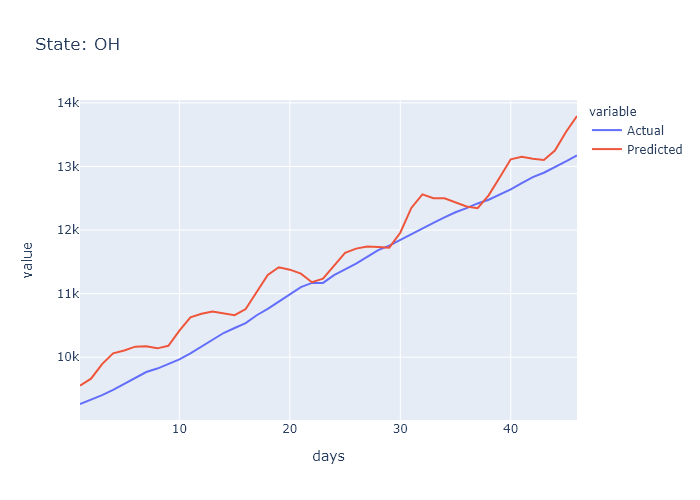

### OK_prediction.png

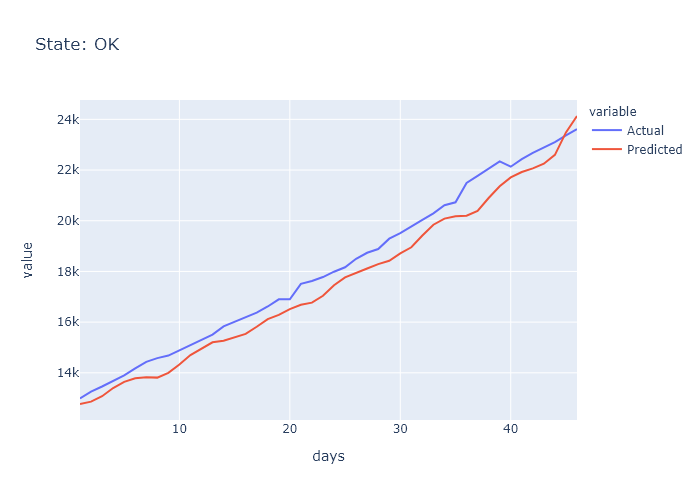

### OK_prediction.png

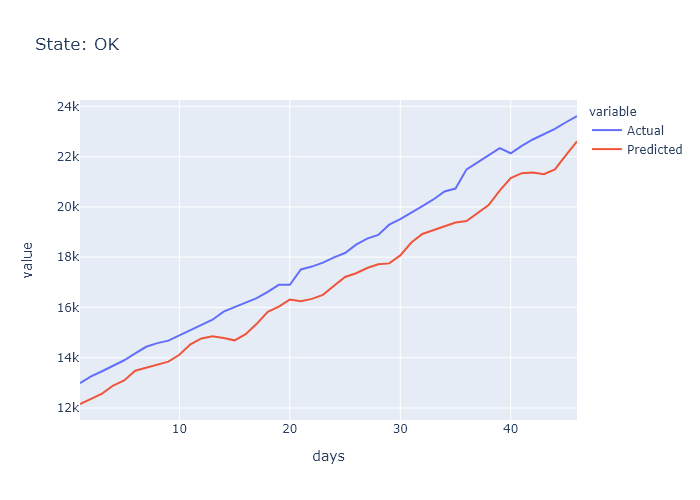

### OR_prediction.png

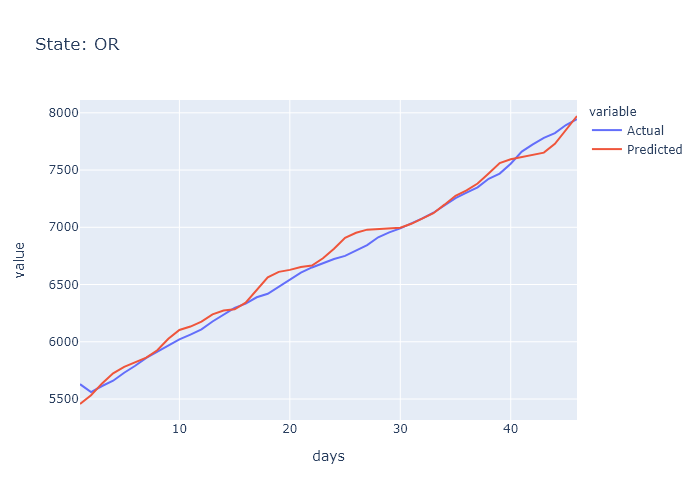

### OR_prediction.png

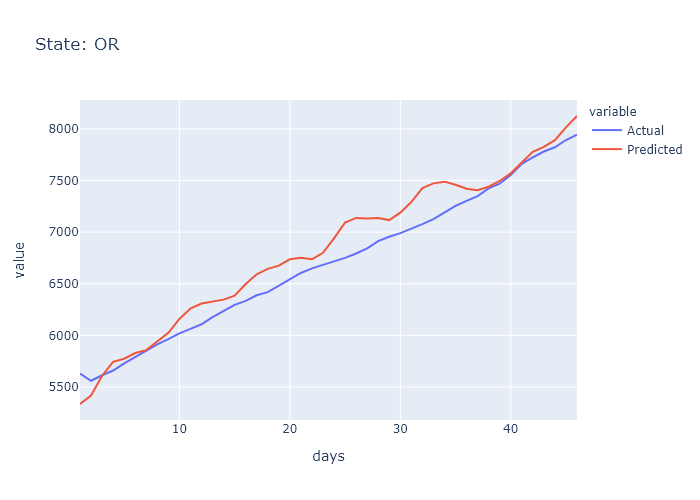

### PA_prediction.png

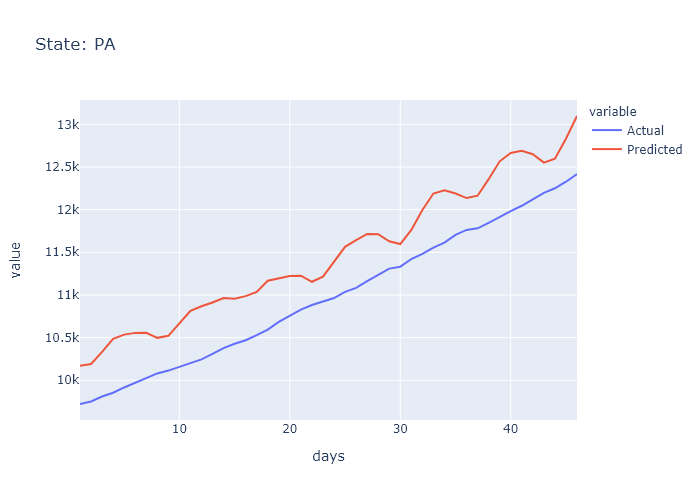

### PA_prediction.png

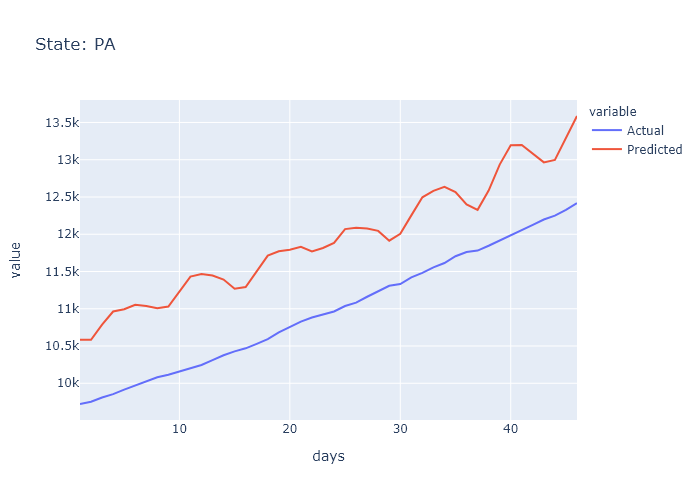

### RI_prediction.png

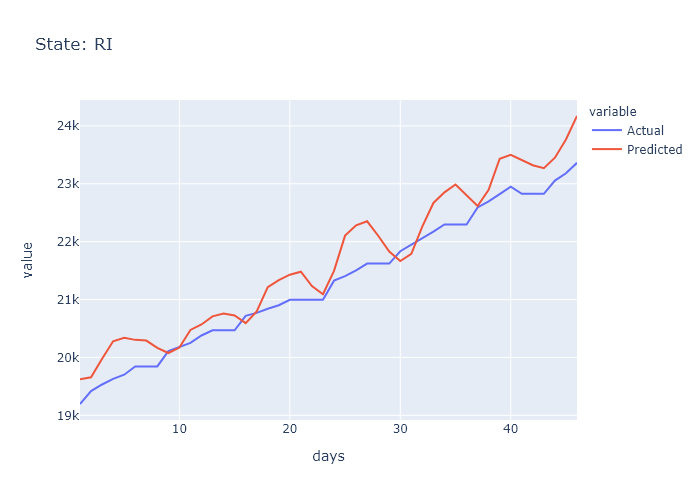

### RI_prediction.png

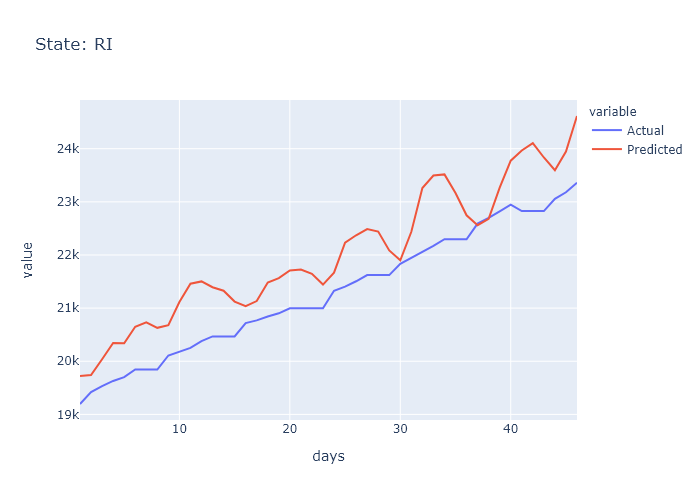

### SC_prediction.png

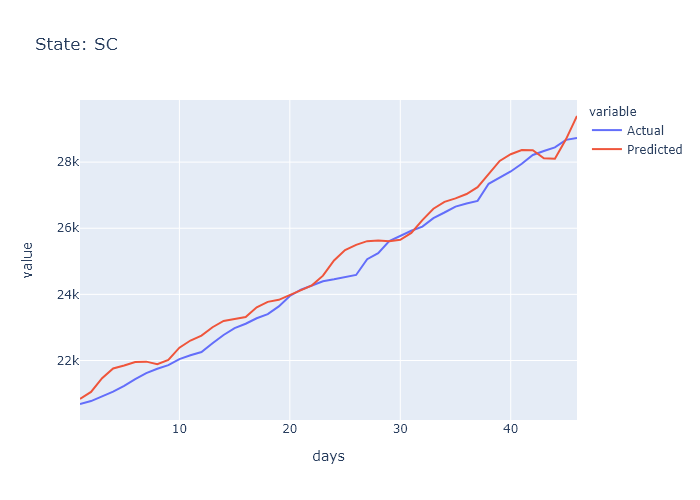

### SC_prediction.png

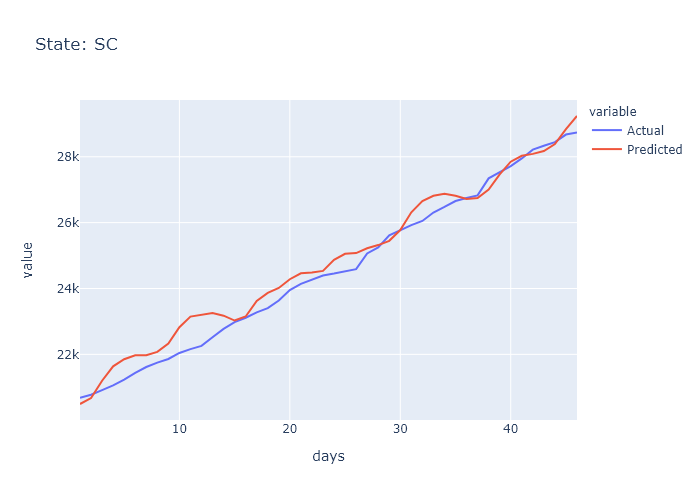

### SD_prediction.png

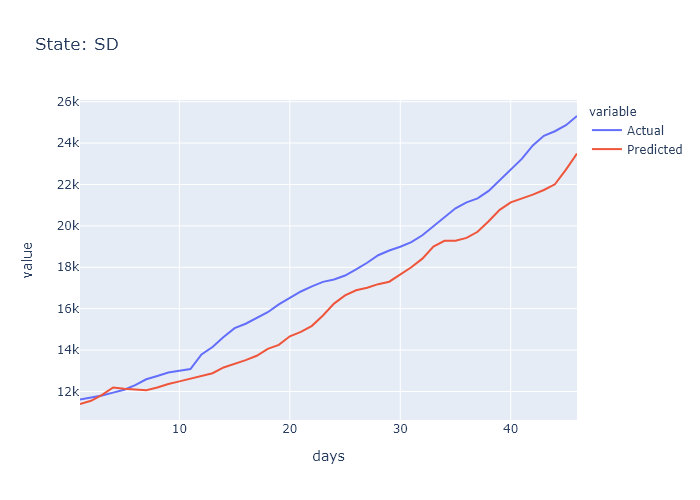

### SD_prediction.png

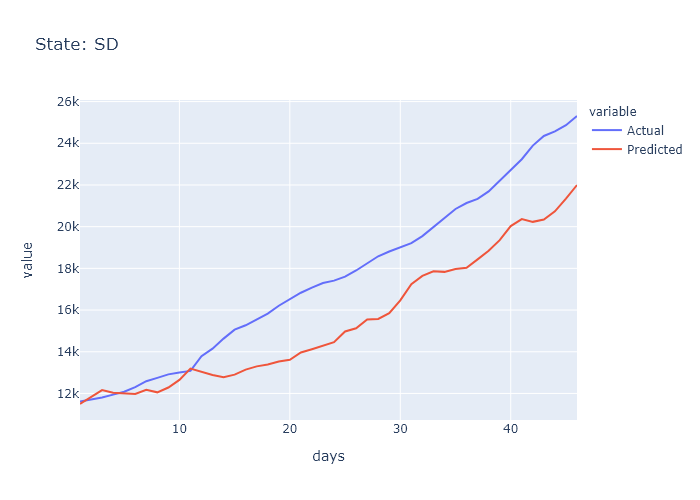

### TN_prediction.png

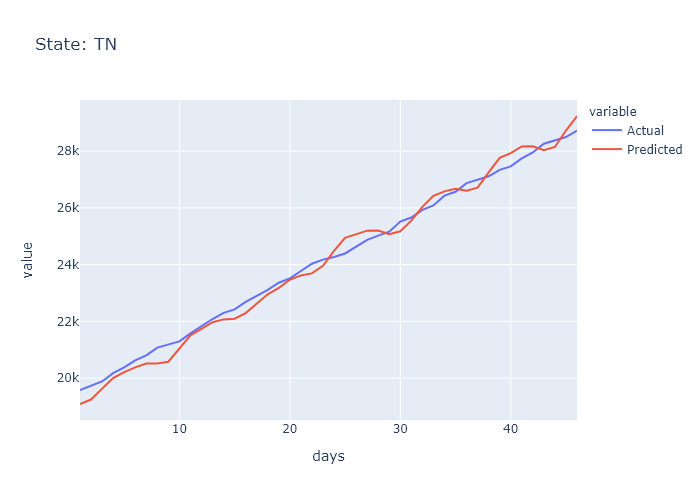

### TN_prediction.png

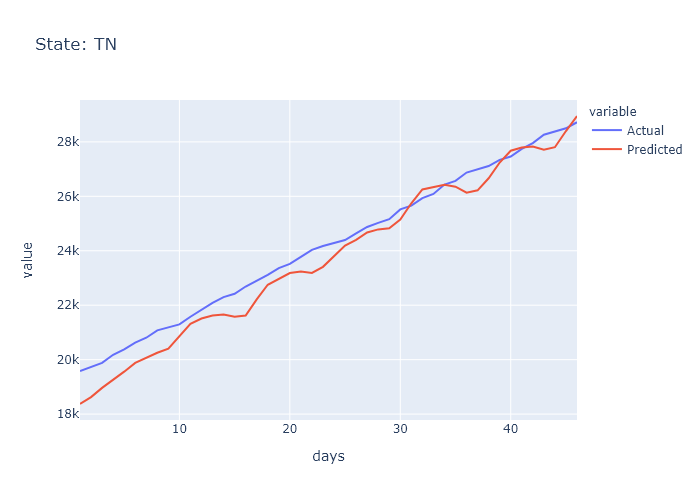

### TX_prediction.png

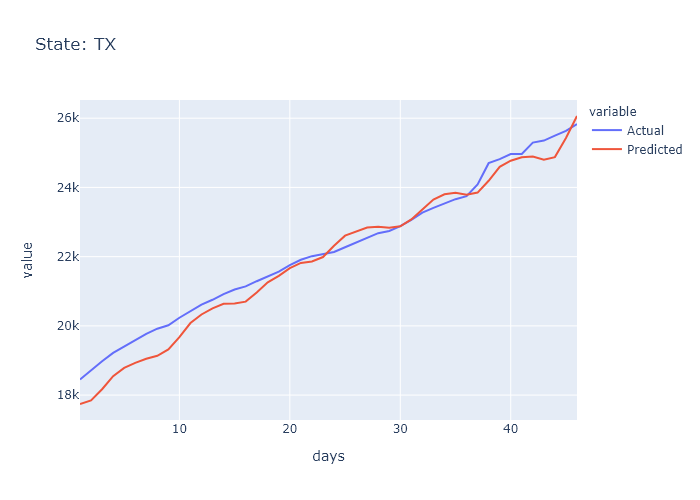

### TX_prediction.png

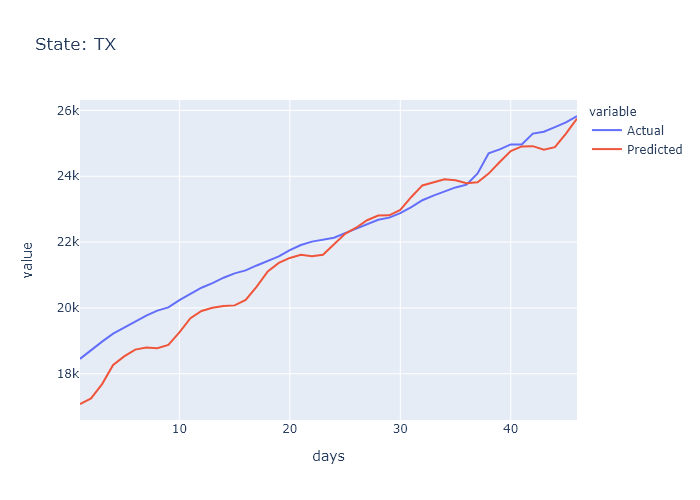

### UT_prediction.png

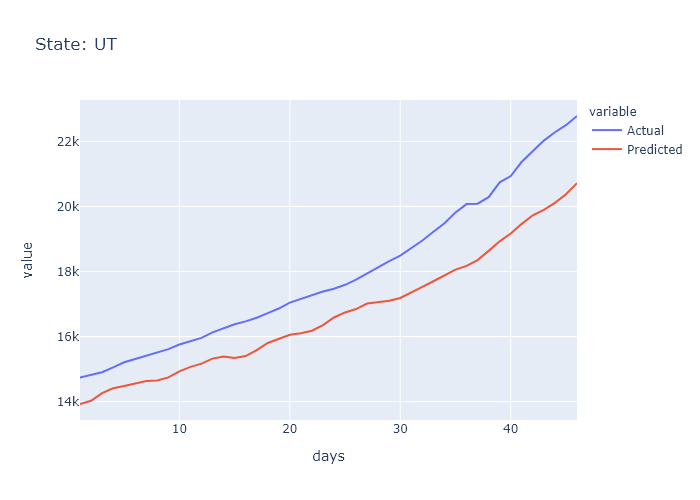

### UT_prediction.png

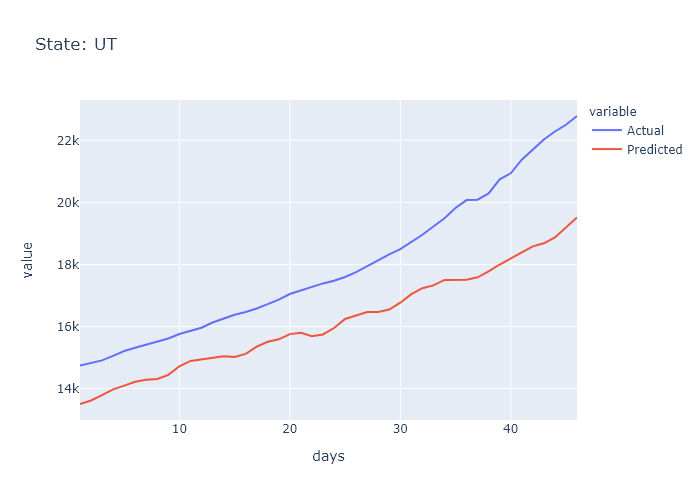

### VA_prediction.png

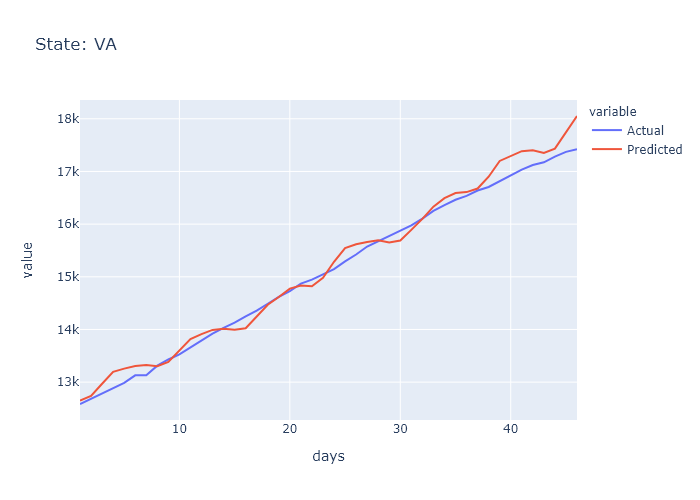

### VA_prediction.png

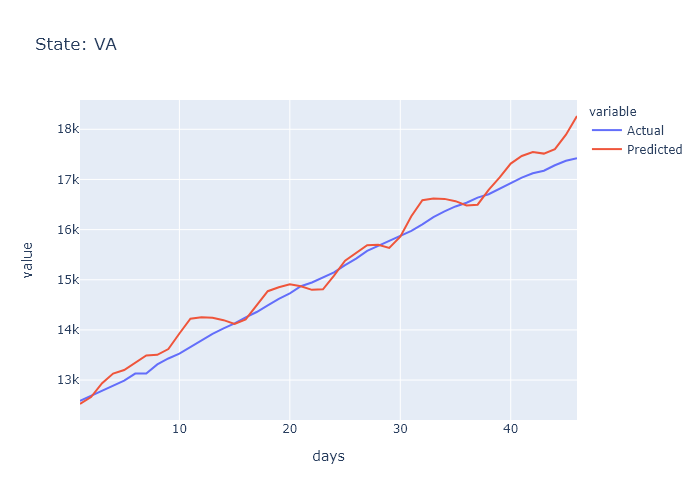

### VT_prediction.png

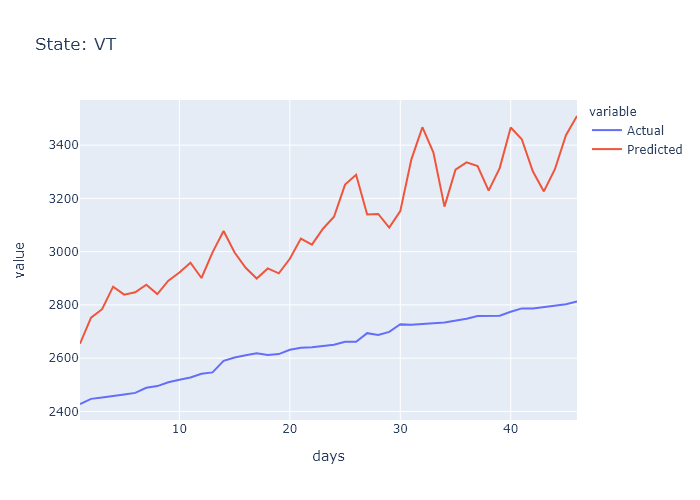

### VT_prediction.png

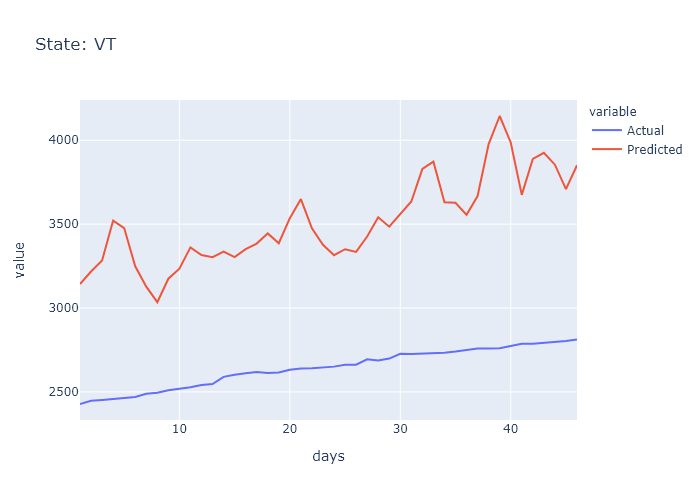

### WA_prediction.png

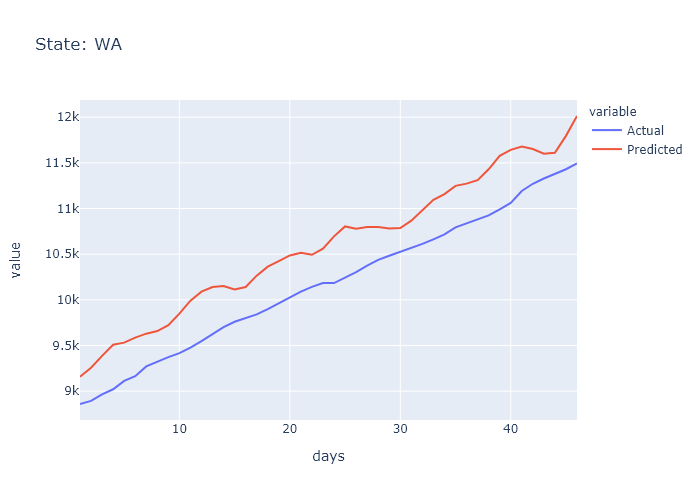

### WA_prediction.png

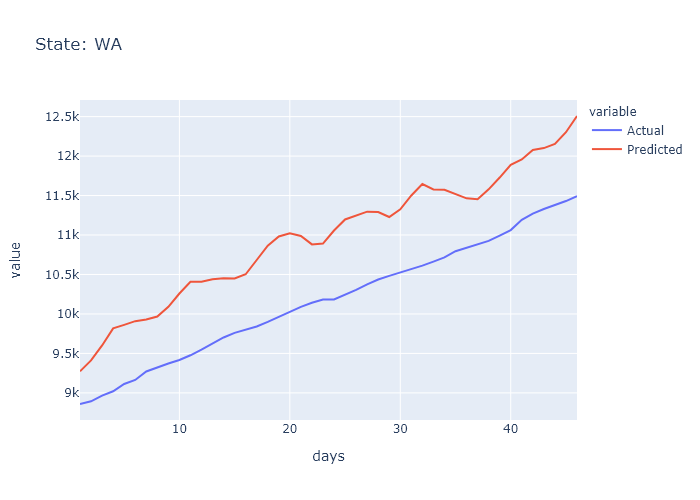

### WI_prediction.png

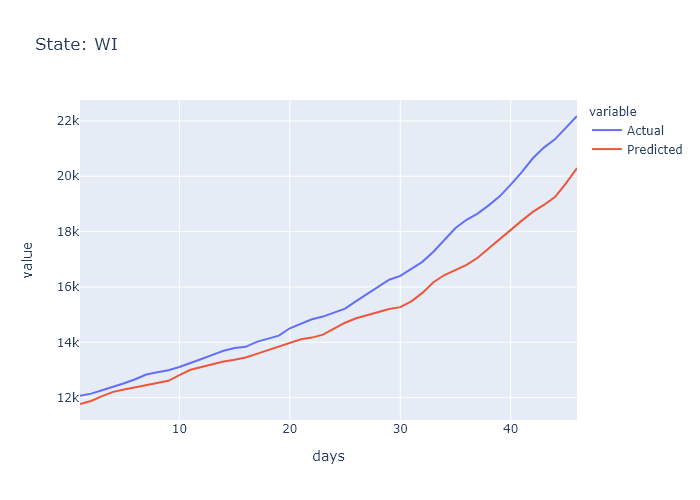

### WI_prediction.png

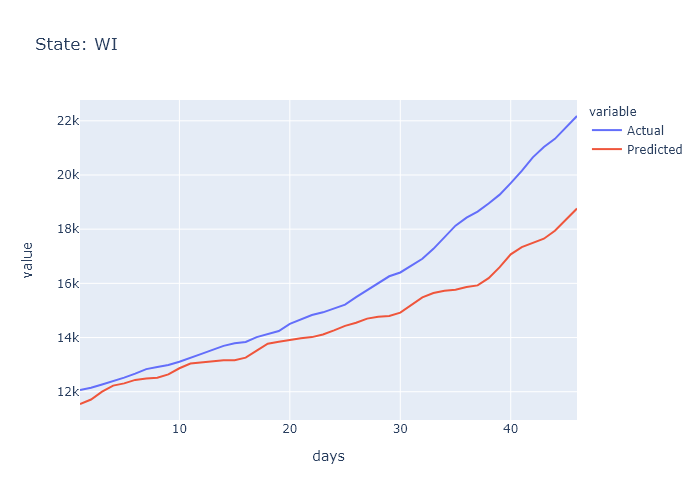

### WV_prediction.png

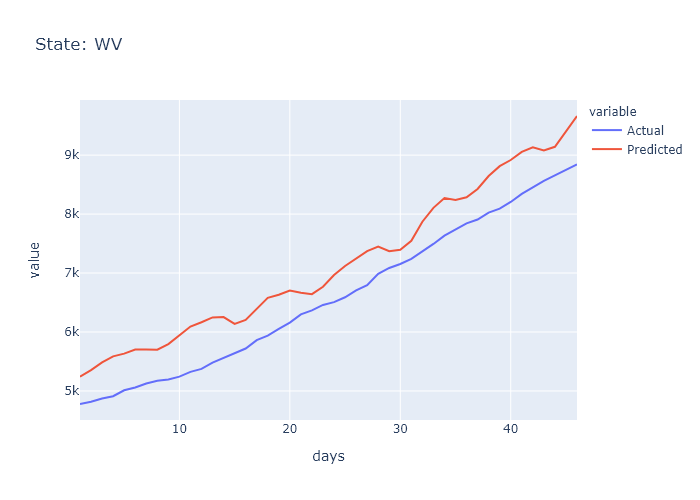

### WY_prediction.png

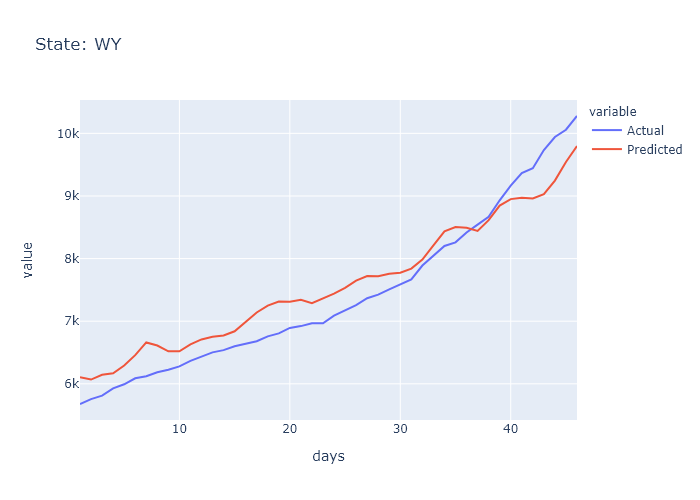
