## Supplementary figures and images for "Improved Prediction of COVID-19 Transmission and Mortality Using Google Search Trends for Symptoms in the United States"

### covid_preds_task1_traj.png

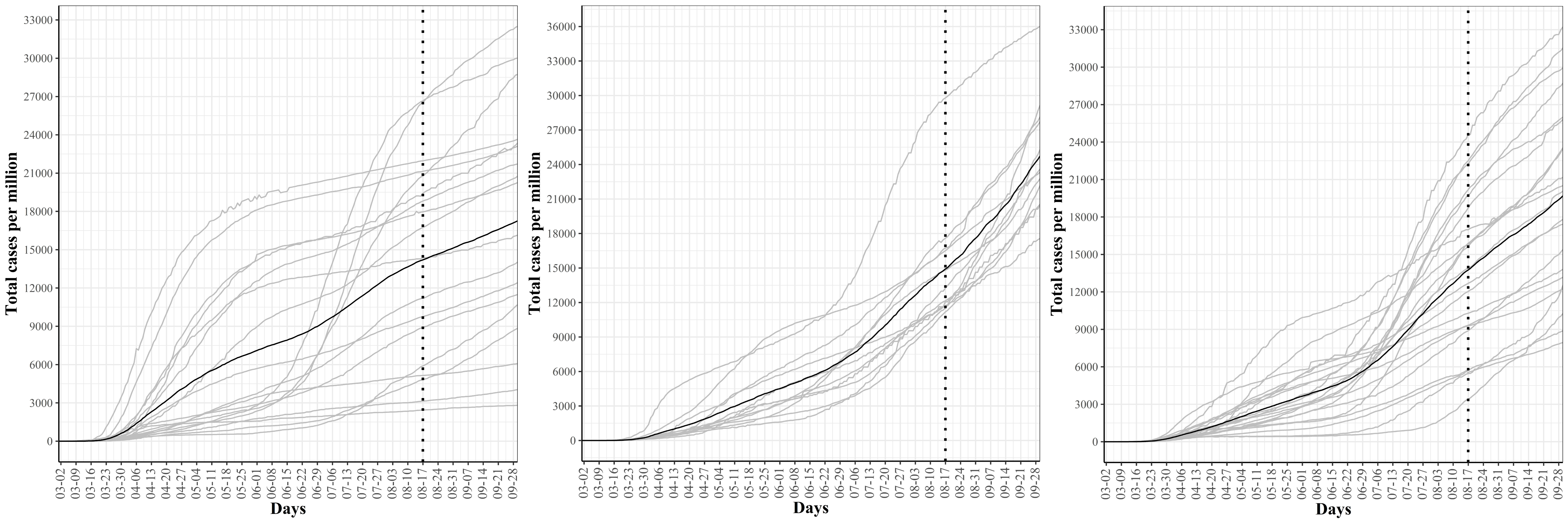

### covid_preds_task2_traj.png

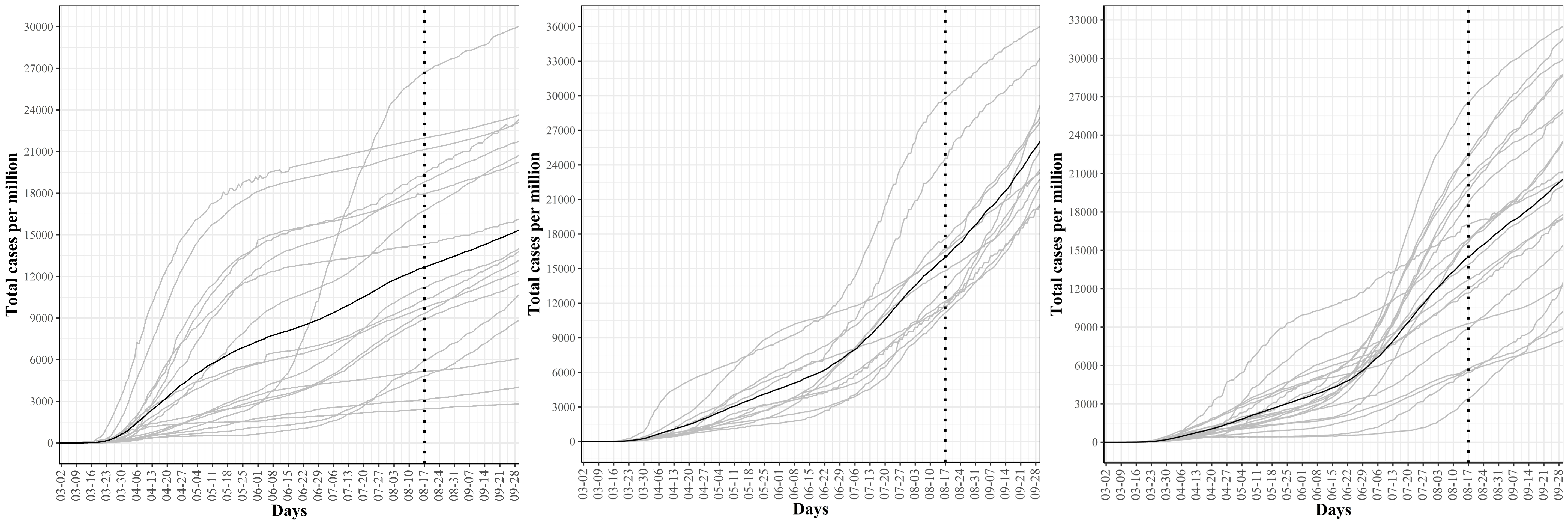

### covid_preds_task3_traj.png

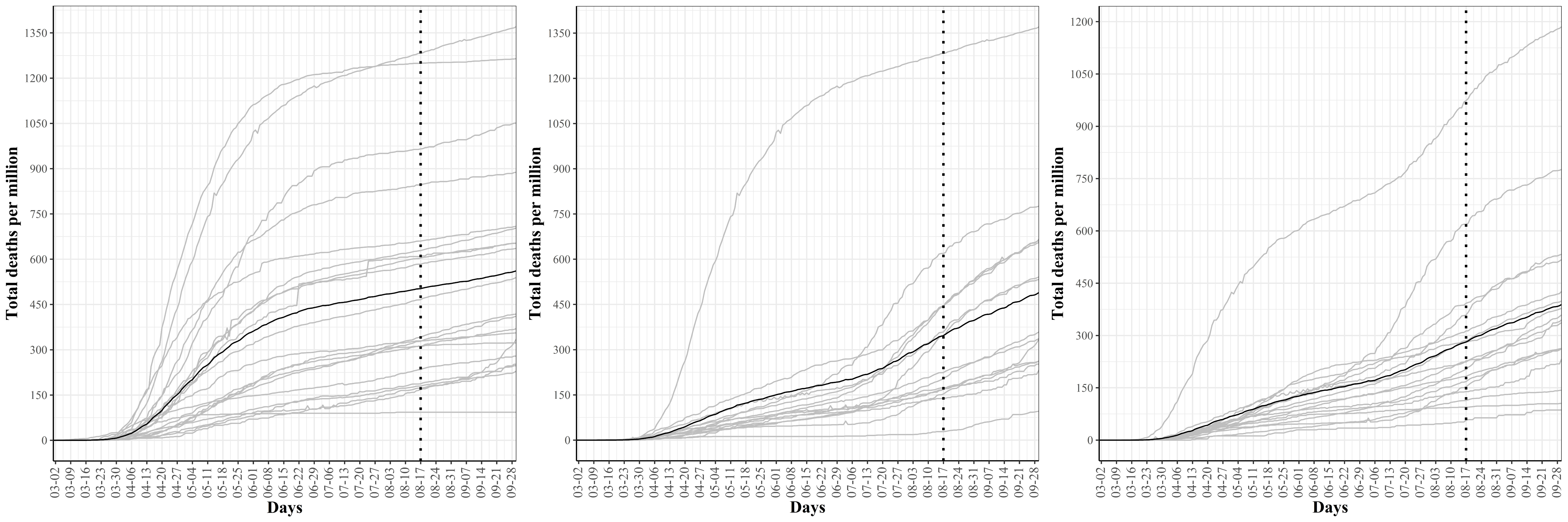

### covid_preds_task4_traj.png

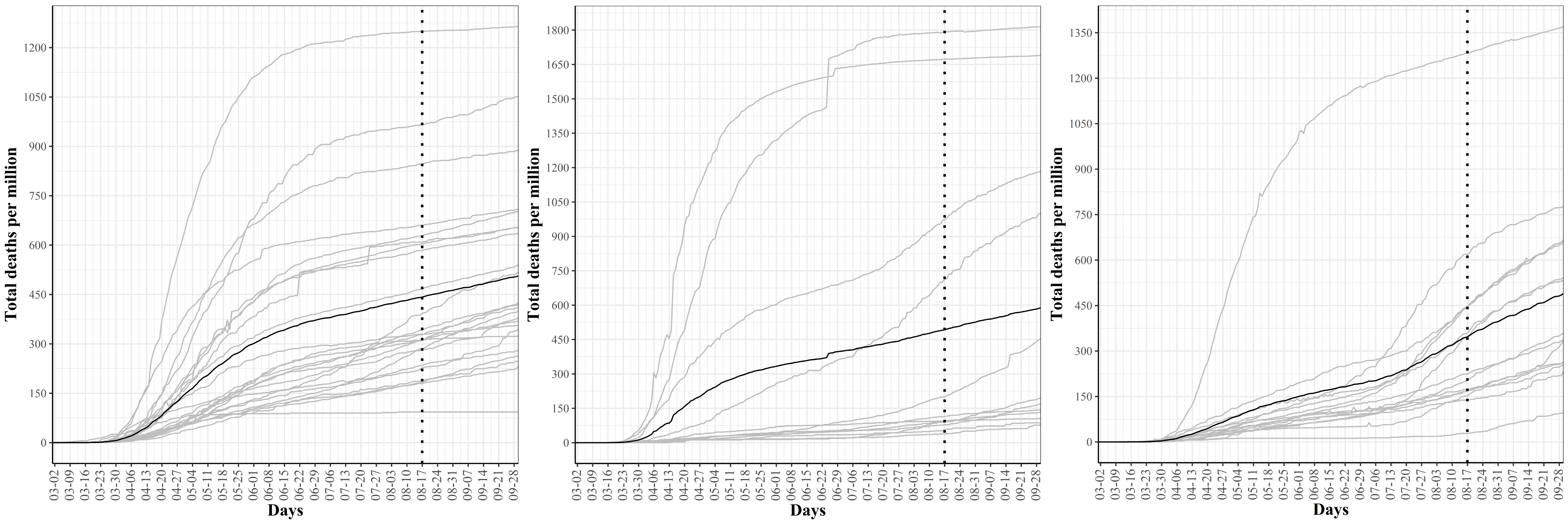

### WA_prediction.png

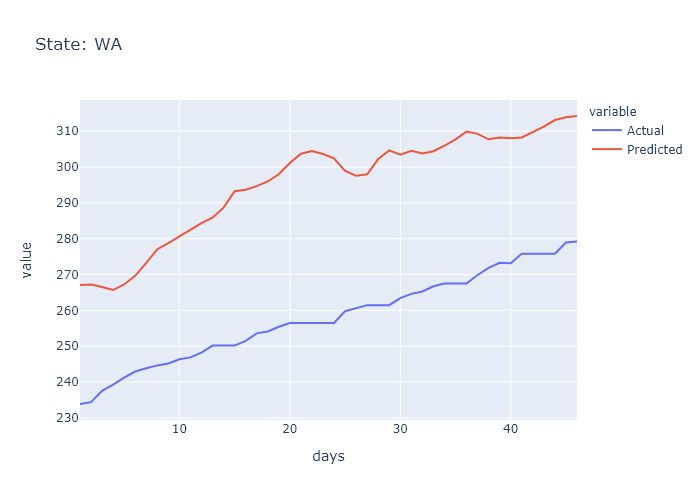

### WI_prediction.png

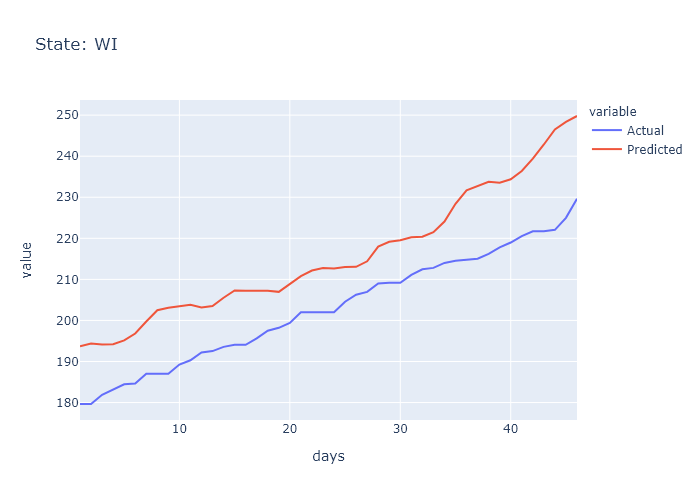

### WV_prediction.png

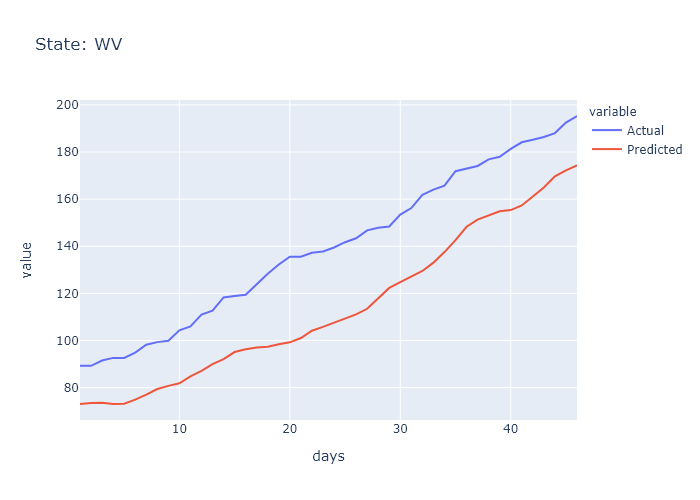

### WY_prediction.png

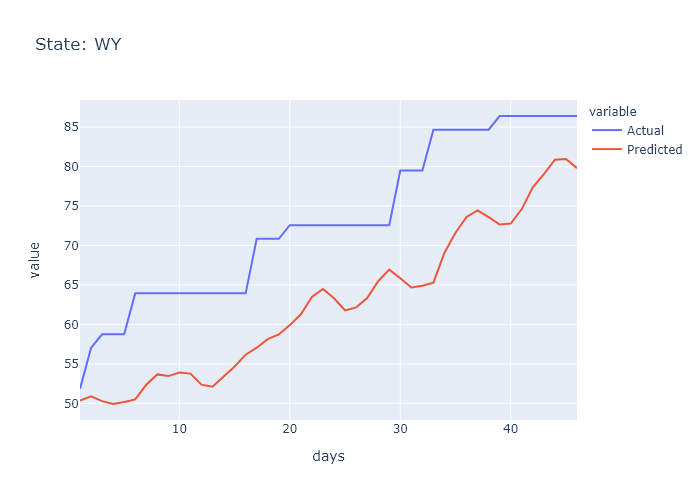
